# An LLM-assisted framework for accelerated and verifiable clinical hypothesis testing from electronic health records

**DOI:** 10.64898/2026.02.10.26346008

**Authors:** Nayoon Gim, In Gim, Yu Jiang, Yuka Kihara, Marian Blazes, Yue Wu, Cecilia S. Lee, Aaron Y. Lee

## Abstract

Acquiring insights from electronic health records (EHRs) is slowed by manual analytical workflows that limit scalability and reproducibility. We present LATCH (LLM-Assisted Testing of Clinical Hypotheses), an agentic framework that converts natural language clinical hypotheses into fully auditable analyses on structured EHR data. LATCH integrates LLM-assisted semantic layers with deterministic execution pipelines to automate cohort construction, statistical analysis, and result reporting, while isolating patient-level data from LLM-involved steps. Using diabetes as a model disease, LATCH reproduced findings from 20 published studies within 3-15 minutes per study. Beyond replication, LATCH enabled study extensions and new insight generation through simple natural language hypothesis modifications. We demonstrated LATCH across 102 hypothesis tests spanning reproduction, extension, and insight generation. We systematically stress-tested LATCH to characterize its limitations and operational boundaries. LATCH provides a scalable framework for reproducible real-world evidence generation, reducing analytical bottlenecks and improving reliability of AI-assisted biomedical discovery while preserving human oversight.

## INTRODUCTION

Despite the growth of data within Electronic Health Records (EHRs), the translation of clinical insights into evidence remains resource-intensive, requiring manual coding for cohort definition, data harmonization, and statistical analysis^1–3^. These labor-intensive steps prolong the research cycle and contribute to the reproducibility crisis, as the exact analytic process is rarely transparent or auditable end-to-end ^4–7^.

Prior attempts to streamline this process have revealed important limitations. Rule-based frameworks, while structured and explainable, often rely on rigid, dataset-specific representations that make it difficult to express nuanced clinical hypotheses, such as defining patient cohorts using combinations of diagnoses and laboratory criteria, and require substantial manual coding effort ^8,9^. There remains a need for a framework that reduces rigidity and coding burden while preserving methodological transparency, enabling efficient hypothesis testing by a broad range of researchers.

While healthcare applications of large language models (LLMs) have focused on tasks such as clinical literature summarization^10,11^, patient engagement^12,13^, or clinical decision support^14,15^, the use of LLMs to support research analytics, translating natural language hypotheses into executable code, remains relatively underexplored. To address this gap, we developed LATCH (Large Language Model-Assisted Testing of Clinical Hypotheses), a framework that automates the technical pipeline from a researcher’s question in natural language to a verifiable statistical analysis. LATCH’s design combines an LLM-integrated semantic layer and a deterministic execution layer. The semantic layer translates natural language hypotheses into explicit cohort definitions and data extraction logic by mapping clinical concepts to the database schema, while statistical analyses and report generation are executed using rule-based procedures to ensure reproducibility.

Prior work has shown that AI approaches operate as opaque systems, limiting verifiability and explainability^16–19^. In contrast, LATCH does not use AI to produce predictions or conclusions. Instead, the LLM is used solely to translate natural language hypotheses into explicit, executable analysis steps, shifting AI output from end results to an analytical process, all of which are fully auditable. Although human review remains essential to ensure methodological rigor, as with any LLM-based application, the analytic report provided by LATCH enables practical human-in-the-loop verification by providing a detailed step-by-step record.

Within the LATCH framework, the researcher remains in control of the scientific inquiry, providing both the hypothesis and the desired analytical method. LATCH translates the query into a series of database operations and statistical analysis by mapping concepts to the database schema and generating the necessary code. LATCH is designed to process structured, tabular health data in relational databases in an LLM vendor agnostic fashion. Importantly, we design LATCH to preserve privacy, only exposing the database schema (e.g., table and column names) to the LLM, while strictly isolating all patient-level data from any LLM-involved steps, addressing privacy concerns regarding LLM use^20–22^.

We demonstrated LATCH’s utility by addressing 102 clinical hypotheses on diabetes, a systemic chronic illness associated with multi-organ complications. Diabetes was selected as a model topic due to its rich foundation of prior publications and publicly available datasets, providing material for investigation across three key scientific use cases: reproducing prior findings, extending existing studies, and generating new insights. All analyses were performed through natural language prompts without requiring any manual coding. LATCH reproduced the findings of 20 published diabetes studies using National Health and Nutrition Examination Survey (NHANES), a dataset of nearly 100,000 patients spanning more than 20 years of survey cycles. Beyond reproduction, LATCH enabled extended analyses on existing studies, including testing cross-dataset generalizability of findings in a different diabetes dataset, AI-READI (Artificial Intelligence Ready and Exploratory Atlas for Diabetes Insights)^23^, testing temporal consistency and performing more granular analyses of NHANES findings. LATCH was also used to generate new insights, identifying a nationwide vision-related trend in the diabetes group from NHANES and subsequently exploring a specific relationship by using the AI-READI cohort to assess the association between disease severity and retinal biomarkers. Lastly, we systematically characterized LATCH’s operational limits and evaluated how built-in safeguards can mitigate, though not eliminate, these situations.

LATCH provides a framework for improving the efficiency of clinical hypothesis testing, supporting the shift of a slow, opaque, and manual process toward a more scalable, accelerated, and transparent workflow. LATCH provides a scalable framework for reproducible real-world evidence generation, reducing analytical bottlenecks and improving reliability of AI-assisted biomedical discovery while preserving human oversight.

## RESULTS

### LATCH: An LLM-assisted framework for verifiable clinical hypothesis testing from EHR data

LATCH is a framework that translates natural language clinical questions into reproducible statistical analyses with structured EHR data. Its architecture strictly isolates LLM-integrated steps from all patient-level operations, ensuring that no patient-level data are exposed to LLMs during any stage of analysis. LATCH is designed to operate on any standardized structured health dataset for which a data dictionary or schema is available. This requires a one-time setup that allows the model to understand the dataset’s variables and their meanings, schema, without accessing patient-level data. This preparation has two components (Extended Data Fig. 1). First, the Code-to-Text Translation step translates coded variable names into natural language descriptions. This not only allows the LLM to accurately interpret variable meaning without prior knowledge of the coding system but also ensures that generated analytic code is interpretable during human review. Second, the Schema Summary Table is constructed to provide an overview of available variables and their attributes. During routine operation, only this schema-level metadata and not patient-level data is shared with the LLM. The resulting schema summary provides the foundation for subsequent schema-grounding steps, enabling LATCH to map natural language concepts to database variables.

LATCH comprises five specialized modules: Planner, Variable Mapper, Data Engine, Statistics Engine, and Reporter (Fig. 1a, Extended Data Fig. 2). These modules are organized into an LLM-assisted semantic layer, which supports code and logic generation from natural language input, and a deterministic execution layer which performs statistical computation and reporting. In this workflow, the Planner converts the clinical question into a structured analysis specification, the Variable Mapper matches this specification to dataset variables using the schema summary, and the Data Engine generates executable database queries to extract the study cohort (Extended Data Fig. 2, Supplementary Fig. 1,2,3). The extracted dataset is processed by the Statistics Engine, and the Reporter produces a comprehensive Analytic Report documenting natural language query, schema mappings, executed code, cohort characteristics, and results (Fig. 1a, Extended Data Fig. 3).

**Fig 1.**
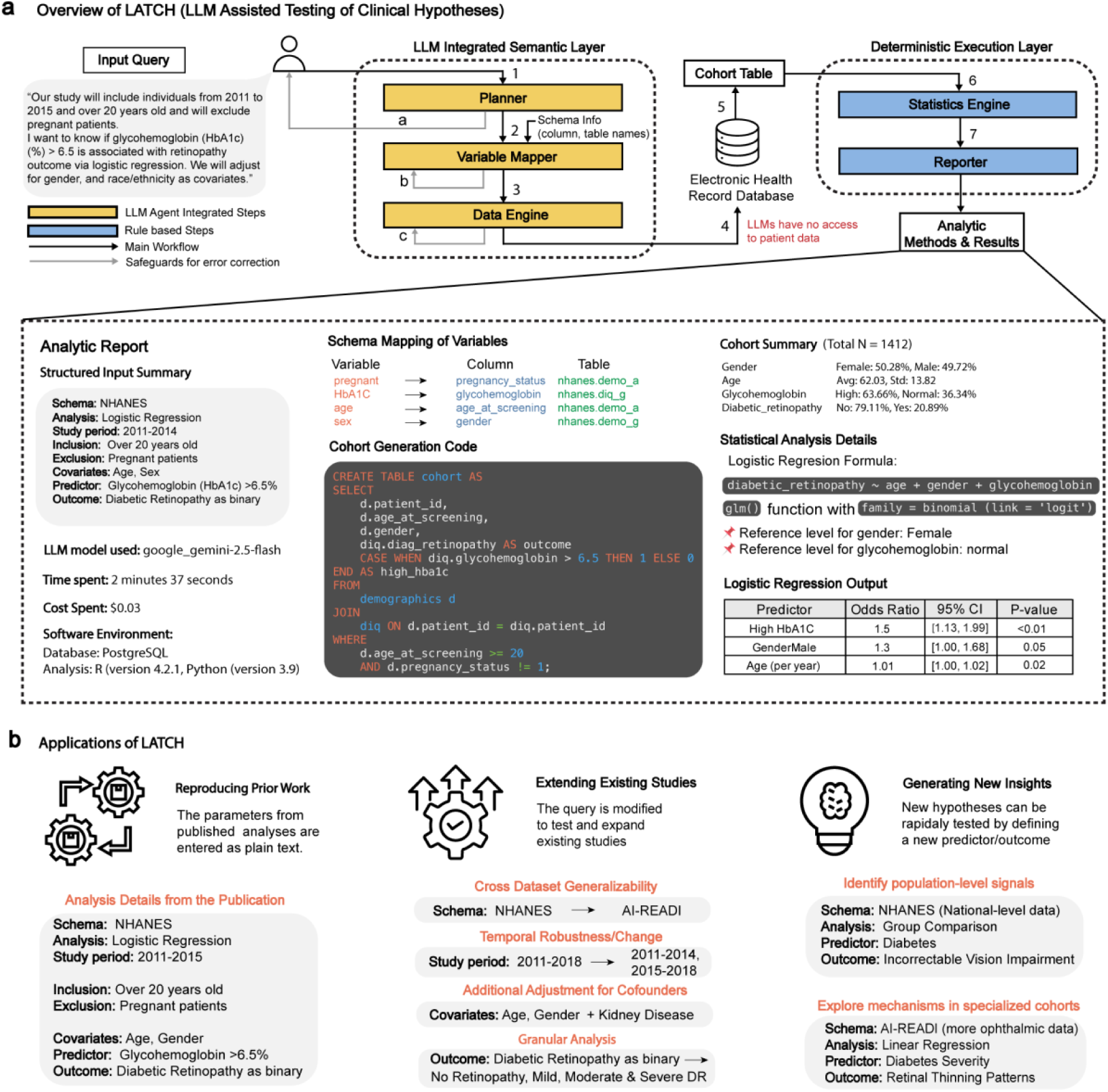
Overview of LATCH framework and its applications. **a,** Architecture and workflow of LATCH. (1) A user’s natural language query is interpreted by the Planner module to generate a structured analysis specification. (2) This specification is passed to the Variable Mapper, which incorporates dataset schema metadata to produce schema-aligned variable mappings. (3) The mapped query is translated into executable database code by the Data Engine to define the study cohort and extract required variables. (4) All cohort extraction and data processing are performed locally, ensuring that patient-level data are never exposed to large language models (LLMs). (5) The extracted dataset is (6) analyzed by a deterministic Statistics Engine. (7) Outputs, including inputs, mappings, executed code, cohort summaries, and statistical results, are recorded by the Reporter module to generate an immutable Analytic Report, enabling transparency and reproducibility. Safeguards are applied across LLM-integrated steps (a-c) to detect and mitigate potential failure modes. **b,** Key applications of LATCH include reproducing published analyses from text-based study specifications (left), extending existing studies through cross-dataset validation, temporal stability assessment, and additional adjustment for confounders, and increased analytical granularity (middle), and generating insights by testing previously unexplored text-based study specifications that define and evaluate new predictors and outcomes (right).

To mitigate errors, LATCH incorporates three safeguards for each of the LLM-integrated steps (Extended Data Fig. 4). First, during the Planner stage, it checks whether the user’s question is missing critical study details or contains internal contradictions. Second, during the Variable Mapping stage, it verifies that selected variables are consistent across survey years and flags any implausible mappings. Third, for the Data Engine, if the generated SQL fails to execute, LATCH automatically repairs and retries up to three times. All analyses are conducted with safeguards enabled, and their impact on error mitigation is evaluated separately, using targeted perturbations of inputs to isolate the effects of each safeguard.

We demonstrate LATCH’s utility across three research applications: reproducing published clinical findings, extending existing studies, and generating new insights. These demonstrations highlight LATCH’s potential to support scalable and reproducible clinical research (Fig.1b).

### Application of LATCH for Reproducing Prior Findings

To evaluate LATCH’s utility, we applied the framework to reproduce the findings of published NHANES-based diabetes studies. Twenty publications were systematically selected based on methodological clarity, citation impact, and relevance to diabetes-related health outcomes. These studies span diverse clinical domains, including diabetic retinopathy^24–30^, diabetic kidney disease^31–35^, mental/cognitive health^36–40^, and all-cause mortality^41–43^, and represent a range of statistical methodologies, including logistic and linear regression, Cox proportional hazards models, and descriptive statistics (Extended Data Fig. 5). For each study, key methodological details from the manuscripts, including inclusion criteria, exclusion criteria, analysis methods, and used variables, were extracted and rephrased into natural language prompt inputs for LATCH (Supplementary Table 1).

Side-by-side comparisons of published results and LATCH-generated outputs are shown in Fig. 2. The concordance between LATCH and published findings was subsequently evaluated using effect direction, statistical significance, and effect size agreement, with summary concordance metrics^44^ shown in Table 1 and Supplementary Table 2.

**Fig 2.**
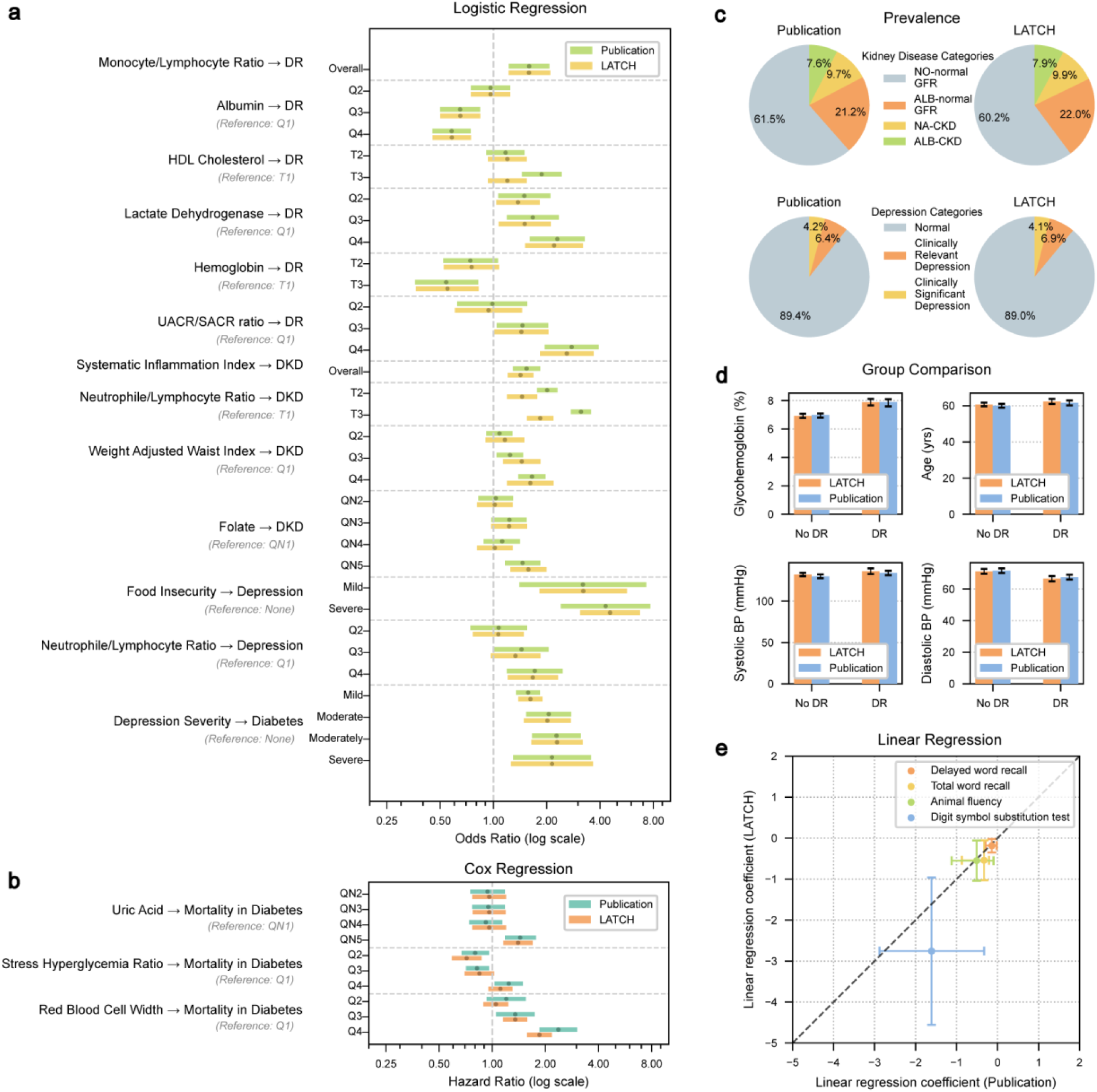
Reproduction of Published Studies. **a,** Logistic regression odds ratios between published results and LATCH results for disease associations. Dots indicate point estimates, with error bars representing 95% confidence intervals (CIs). **b,** Cox regression hazard ratios between published results and LATCH results for all-cause mortality-related studies. Dots indicate point estimates, with error bars representing 95% CIs. **c,** Prevalence of different kidney disease types and depression severity in diabetes between published results. **d,** Group comparisons between patients with and without diabetic retinopathy ratios between published results and LATCH results. Error bars indicate 95% CIs. **e,** Linear regression coefficients for the effect of glycohemoglobin on cognitive scores as reported in published results (x axis) versus those calculated by LATCH (y axis). Dots indicate point estimates, with error bars representing 95% CIs. Abbreviations: QN, quintile; Q, quartile; T, tertile; DR, diabetic retinopathy; DKD, diabetic kidney disease; HDL, high-density lipoprotein; UACR/SACR, urinary serum albumin-to-creatinine ratio; NA-normal GFR, normal albuminuria normal glomerular filtration rate; ALB-normal GFR, albuminuria with normal glomerular filtration rate; NA-CKD, normal albuminuria chronic kidney disease defined by glomerular filtration rate; ALB-CKD, albuminuria with chronic kidney disease. Concordance evaluation results across different metrics are provided in Table 1 and Supplementary Table 2.

**Table 1.** Summary of the concordance evaluation from the reproduction study. Concordance was assessed across multiple metrics, including effect direction (whether both analyses indicated increased or decreased risk), p-value concordance (whether both results were statistically significant at p < 0.05 or both were non-significant), and overlap of 95% confidence intervals (CIs). A check mark indicates concordance, whereas a cross indicates discordance. Additional descriptive measures were used to support interpretation, including comparison of analytic sample sizes between the LATCH reproduction and the original published studies to assess cohort alignment, and calculation of relative effect size ratios (published estimate divided by the LATCH estimate). P-value concordance, confidence interval overlap, and relative effect size comparisons were based on the most extreme exposure level (for example, highest quartile or highest tertile). This table presents concordance results for logistic and Cox regression analyses, which constituted the majority of reproduced models, and is limited to these models because other analysis types reported heterogeneous outcome metrics that could not be consistently summarized within a single table. The reported time reflects the elapsed duration from query initiation to report of final results. Reported runtimes are approximate and may vary with network conditions and token usage, which can differ slightly across runs even for identical queries. Additional information for all 20 studies, including references to the original publications and Figure 2, are provided in Supplementary Table 2.

| Analysis | Regression Type | Sample Size (Pub / LATCH) | Effect Direction Concordant | P value Concordant | CI Overlap | Relative Effect (Pub / LATCH) | Time (min) |
| --- | --- | --- | --- | --- | --- | --- | --- |
| Monocyte to Lymphocyte Ratio → Diabetic Retinopathy | Logistic | 367 / 367 | ✓ | ✓ | ✓ | 1.00 | 3.1 |
| Albumin → Diabetic Retinopathy | Logistic | 2,964 / 2,964 | ✓ | ✓ | ✓ | 1.00 | 4.0 |
| HDL Cholesterol → Diabetic Retinopathy | Logistic | 1,708 / 1,380 | ✓ | ✗ | ✓ | 1.56 | 3.4 |
| Lactate Dehydrogenase → Diabetic Retinopathy | Logistic | 3,476 / 3,476 | ✓ | ✓ | ✓ | 1.04 | 3.8 |
| Hemoglobin → Diabetic Retinopathy | Logistic | 837 / 844 | ✓ | ✓ | ✓ | 0.98 | 1.6 |
| UACR/SACR Ratio → Diabetic Retinopathy | Logistic | 2,594 / 2,594 | ✓ | ✓ | ✓ | 1.07 | 5.7 |
| Systematic Inflammation Index → Diabetic Kidney Disease | Logistic | 3,937 / 3,822 | ✓ | ✓ | ✓ | 1.08 | 6.5 |
| Neutrophil to Lymphocyte Ratio → Diabetic Kidney Disease | Logistic | 7,153 / 7,048 | ✓ | ✓ | ✗ | 1.70 | 15.1 |
| Weight Adjusted Waist Index → Diabetic Kidney Disease | Logistic | 5,028 / 5,155 | ✓ | ✓ | ✓ | 1.02 | 8.0 |
| Folate → Diabetic Kidney Disease | Logistic | 3,461 / 3,396 | ✓ | ✓ | ✓ | 0.93 | 4.9 |
| Food Insecurity → Depression in Diabetes | Logistic | 1,724 / 1,688 | ✓ | ✓ | ✓ | 0.94 | 3.7 |
| Neutrophil to Lymphocyte Ratio → Depression in Diabetes | Logistic | 2,820 / 2,703 | ✓ | ✓ | ✓ | 1.03 | 8.7 |
| Depression Severity → Diabetes | Logistic | 14,328 / 14,418 | ✓ | ✓ | ✓ | 1.00 | 8.4 |
| Uric Acid → Mortality in Diabetes | Cox | 7,101 / 6,832 | ✓ | ✓ | ✓ | 1.03 | 14.2 |
| Stress Hyperglycemia Ratio → Mortality in Diabetes | Cox | 11,160 / 10,827 | ✓ | ✗ | ✓ | 1.12 | 8.3 |
| Red Blood Cell Width → Mortality in Diabetes | Cox | 3,061 / 3,157 | ✓ | ✓ | ✓ | 1.28 | 5.9 |

In logistic regression analyses (Fig. 2a), LATCH reproduced both the direction and magnitude of many published odds ratios (ORs) for diabetic retinopathy. For example, the association between monocyte-to-lymphocyte ratio and diabetic retinopathy was closely matched (published OR = 1.59 (*n* = 367), 95% confidence interval (CI): 1.22–2.07; LATCH OR = 1.59 (*n* = 367), 95% CI: 1.22–2.09). The LATCH result reported a more precise P value that was rounded in the original publication (published *P* ≤ 0.001; LATCH *P* ≤ 7.94 × 10^−4^). LATCH also reproduced protective associations of serum albumin and hemoglobin with diabetic retinopathy, as well as reported risk gradients for diabetic kidney disease and depression across various risk factors (Fig. 2a). In Cox proportional hazards models (Fig. 2b), LATCH identified elevated all-cause mortality risk among patients with diabetes associated with higher uric acid and red blood cell distribution width. Beyond regression, prevalence estimates (Fig. 2c), descriptive comparisons of numerical variables (Fig. 2d), and linear regression analyses (Fig. 2e) were reproduced. Reconstructed cohort sizes matched those reported in the original studies, with most differing by less than 10% and several matching exactly (Table 1). End-to-end reproductions were completed efficiently, with runtimes ranging from 3 to 15 minutes.

Three discrepancies were examined (Table 1). In the HDL cholesterol-retinopathy study, the original study reported significance for the highest tertile (T3), which was not observed with LATCH. This discrepancy likely resulted from ambiguity in the diabetes inclusion criteria, which combined “OR” conditions with comma-separated clauses, leading to a different logical interpretation and a smaller cohort (publication *n =* 1,708 vs. LATCH *n =* 1,380). In the neutrophil-to-lymphocyte ratio and diabetic kidney disease analysis, both found significant associations, but with non-overlapping confidence intervals. More than one NHANES variable could represent pregnancy and cancer exclusions or kidney disease-related laboratory measures, and our implementation may have selected similarly labeled fields. For the stress hyperglycemia ratio-mortality analysis, NHANES lacks a direct analogue to “admission glucose,” and our choice of proxy could have differed from the original’s. Although these factors provide plausible explanations based on available documentation and data constraints, the absence of access to original analysis code precludes definitive attribution of the underlying causes. Overall, effect size differences likely reflect minor variation in cohort construction and variable interpretation, along with common reproducibility challenges such as ambiguous cohort definitions, undocumented preprocessing decisions, and software implementation differences.

To evaluate the stability and reproducibility of LATCH-generated analyses given the non-deterministic nature of large language models, we conducted repeated reproductions (*n=* 3) of 20 studies (Extended Data Fig. 6). LATCH produced consistent outputs in most cases, with 17 studies fully concordant and 3 showing variation, primarily due to reasonable but different methodological interpretation, and one case resulting from misimplementation. For these cases, Fig. 2 shows the most frequent output. Together, these findings highlight the importance of human oversight to ensure system-generated analyses align with the intended analysis.

### Application of LATCH for Extending of Existing Studies

We used LATCH to extend prior NHANES-based studies by evaluating cross-dataset generalizability, temporal consistency, potential confounding and outcome granularity all through modifications to natural language prompts.

To assess cross-dataset generalizability (Fig. 3a), we modified the prompt to use the AI-READI dataset instead of NHANES. We evaluated three diabetic retinopathy risk factors reproduced from NHANES-based studies and ten additional risk factors mentioned in the American Academy of Ophthalmology (AAO) Diabetic Retinopathy Preferred Practice Pattern^45^ across both NHANES and AI-READI datasets. A high urinary albumin-to-creatinine ratio/ serum albumin-to-creatinine ratio (UACR/SACR) ratio (Q4) (AI-READI adjusted *P* ≤ 1.57 × 10^−4^; NHANES adjusted *P* ≤ 4.21 × 10^−10^), HbA1c ≥ 7.0% (AI-READI adjusted *P* ≤ 1.76 × 10^−2^; NHANES adjusted *P* ≤ 4.79 × 10^−12^), diabetes duration ≥ 10 years (AI-READI adjusted *P* ≤ 3.33 × 10^−4^; NHANES adjusted *P* ≤ 4.07 × 10^−32^), kidney disease presence (AI-READI adjusted *P* ≤ 1.56 × 10^−3^; NHANES adjusted *P* ≤ 1.09 × 10^−32^), and insulin use (AI-READI adjusted *P* ≤ 2.46 × 10^−9^; NHANES adjusted *P* ≤ 1.62 × 10^−64^) remained significantly associated with diabetic retinopathy in both datasets, whereas a high serum albumin (Q4) (NHANES adjusted *P* ≤ 1.22 × 10^−3^) and hypertensive systolic blood pressure (NHANES adjusted *P* ≤ 6.64× 10^−3^) were significantly associated only in NHANES.

**Fig 3.**
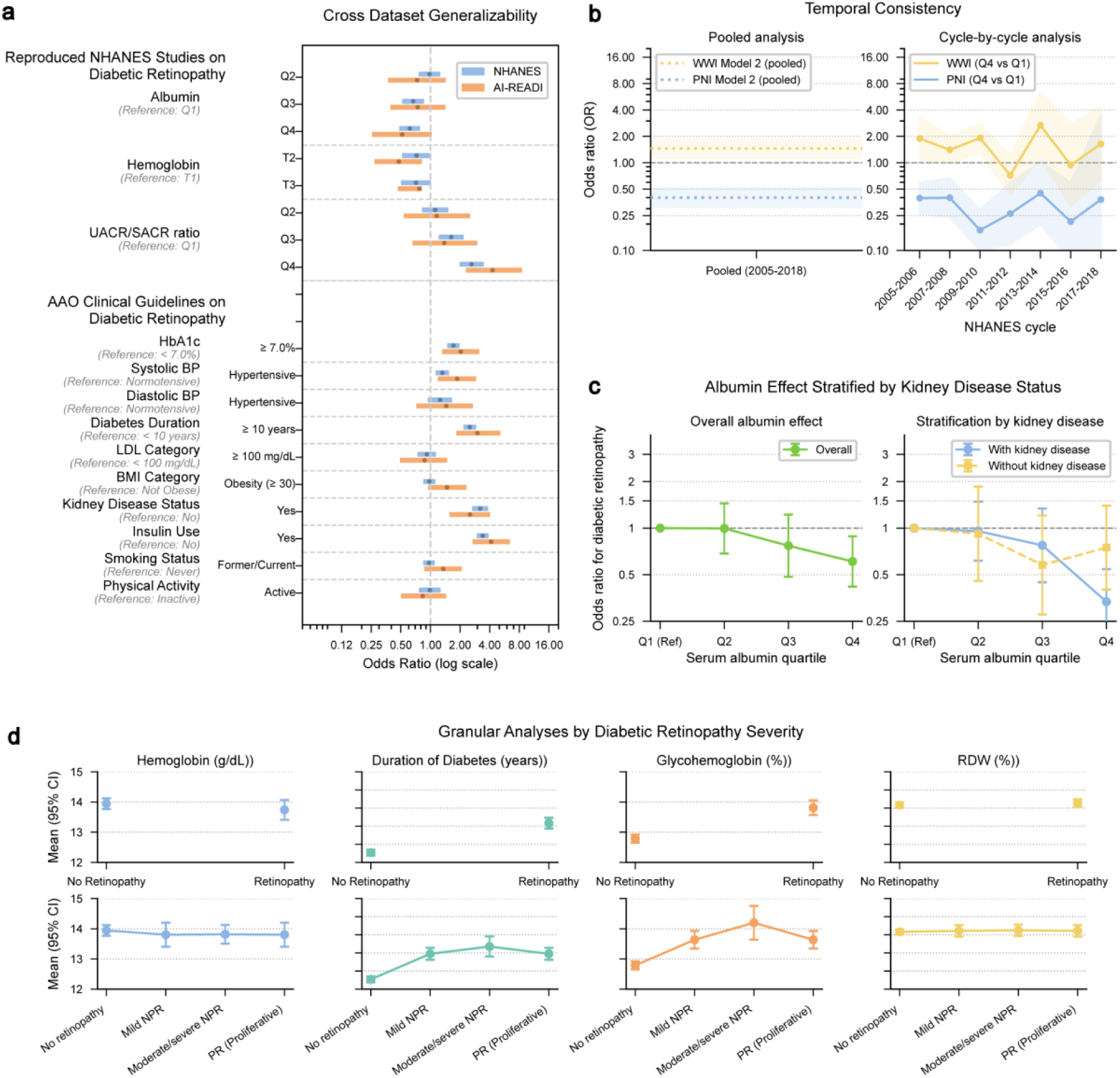
Extension of Existing Studies. **a,** Cross-dataset generalizability. Forest plot comparing odds ratios (ORs) for established diabetic retinopathy (DR) risk factors across the NHANES (blue) and AI-READI (orange) cohorts. Variables include risk factors reproduced from prior NHANES-based studies as well as those in the American Academy of Ophthalmology (AAO) Diabetic Retinopathy Preferred Practice Pattern. Dots indicate point estimates and error bars represent 95% confidence intervals (CIs). **b,** Temporal consistency. Associations between prognostic nutritional index (PNI) and mortality, and between weight-adjusted waist index (WWI) and mortality, shown for pooled analyses (left) and across individual NHANES survey cycles from 2005-2006 through 2017-2018 (right). Dots indicate point estimates and shaded areas represent 95% CIs. **c,** Stratified confounder-adjusted analysis by kidney disease status. Odds ratios for DR across serum albumin quartiles shown without stratification (left) and stratified by kidney disease status (right). Dots indicate point estimates, with error bars representing 95% CIs. **d,** Granular analyses of laboratory values by diabetic retinopathy (DR) severity. The top panels replicate the original binary groups from prior studies, comparing the presence versus absence of DR. The bottom panels extend this analysis using LATCH by categorizing DR into four ordered severity levels, enabling finer-grained assessment of biomarker trends across disease progression. Dots indicate point estimates, with error bars representing 95% CIs.

To evaluate temporal consistency (Fig. 3b), we adjusted the prompt to analyze seven NHANES cycles rather than a single study period. The Prognostic Nutritional Index (PNI)^46^ demonstrated a consistently protective association across all survey cycles. In contrast, the Weight-Adjusted Waist Index (WWI)^47^ exhibited substantial temporal variability, including direction reversals in the 2011–2012 cycle (OR = 0.72 (*n* = 872), 95% CI: 0.43, 1.20, *P* ≤ 2.08 × 10^−1^) and the 2015–2016 cycle (OR = 0.94 (*n* = 1028), 95% CI: 0.32, 2.79, *P* ≤ 9.16 × 10^−1^), where odds ratios fell on opposite sides of the null value (OR = 1) compared with other cycles. Despite this heterogeneity, the pooled analysis indicated a significantly elevated risk associated with WWI.

Single-factor analyses may yield associations without sufficient clinical context^48^. To address this limitation, we used LATCH to reevaluate the reported association between low serum albumin and diabetic retinopathy^26^ while accounting for an additional confounding variable, chronic kidney disease (CKD), a known cause of hypoalbuminemia and an established retinopathy risk factor ^49,50^ (Fig. 3c). After stratifying by CKD status, a strong association was observed among individuals with CKD (OR = 0.33, 95% CI: 0.21, 0.54, *P* ≤ 2.80 × 10^−5^), but not among those without CKD (OR = 0.75; 95% CI: 0.40, 1.40, *P* ≤ 3.58 × 10^−1^). These results indicate that the overall association was largely driven by participants with CKD.

Lastly, we extended the hemoglobin-based analysis^29^ by substituting its binary retinopathy outcome with a four-level retinopathy severity scale, allowing more granular characterization of biomarker trends across disease progression (Fig. 3d).

### Application of LATCH for Generating New Insights

Beyond replicating prior studies, we used LATCH to generate new insights through a sequential, hypothesis-driven workflow. Focusing on diabetes and vision outcomes, we used LATCH to execute analyses expressed in natural language, with each analysis motivated by the previous one. We first identified population-level patterns in diabetes prevalence across vision impairment groups in NHANES, then extended these findings to the AI-READI cohort, which provides richer vision phenotyping and retinal structural measures (Fig. 4a). This iterative process was enabled by LATCH’s ability to reduce technical overhead while preserving transparency and analytical control.

**Fig 4.**
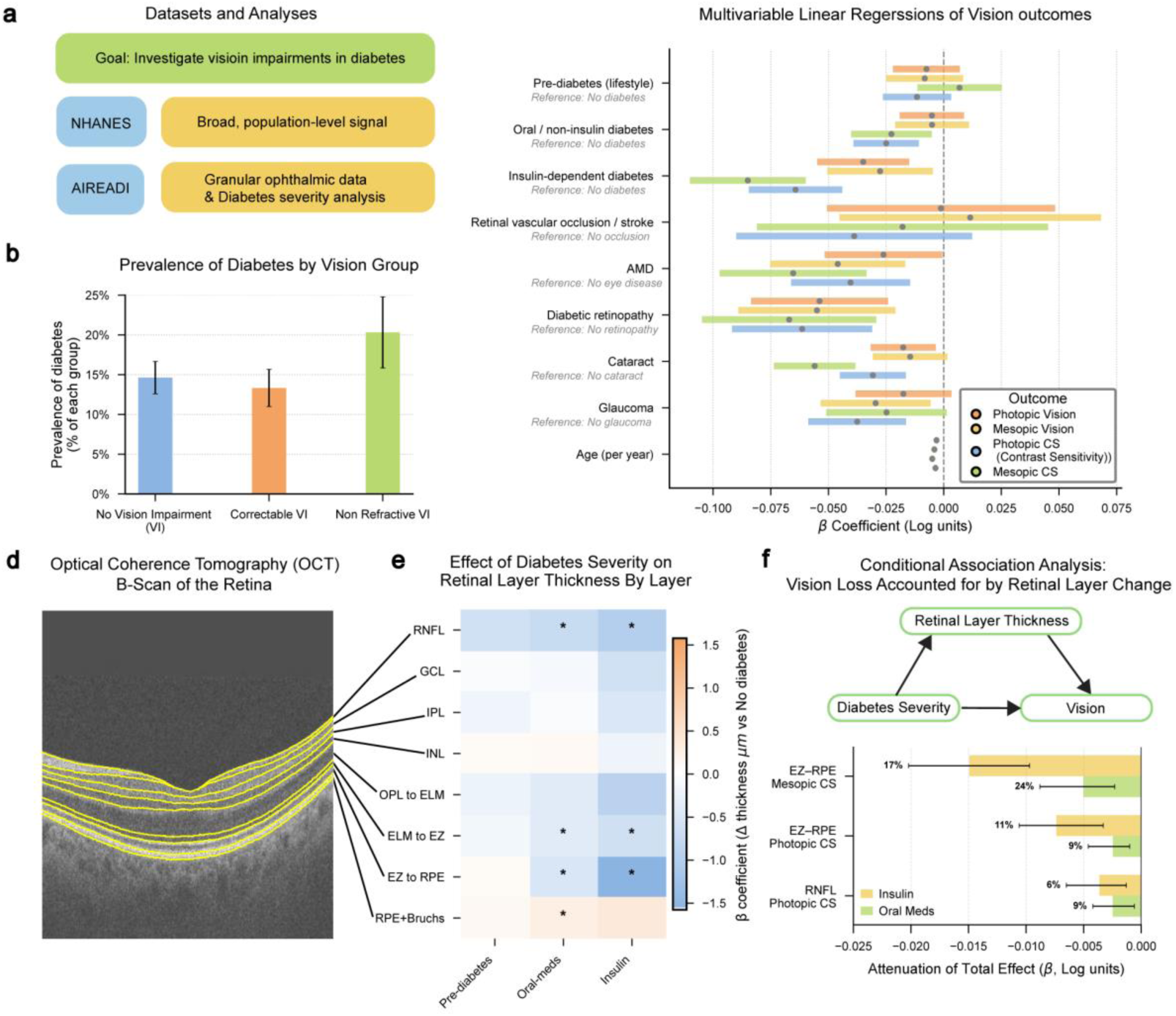
Generation of New Insights. **a,** A schematic illustrating population-level analyses, followed by granular ophthalmic phenotyping to investigate visual impairments in diabetes. **b,** Population-Level Association. Analysis reveals a significant difference in diabetes prevalence across visual impairment groups: no vision impairment (VI), correctable VI, and non-refractive (uncorrectable) VI (*P* ≤ 2.91 × 10^−2^) from a survey-weighted group comparison using the Rao–Scott chi-square test. The highest prevalence of diabetes was observed among individuals with non-refractive VI. Error bars indicate 95% confidence intervals (CIs). **c,** Visual Function Analysis. linear regression coefficients from four separate models assessing the association between diabetes severity and visual function outcomes, including photopic and mesopic vision (−logMAR) and contrast sensitivity (log). Larger coefficients indicate better visual performance. Dots indicate point estimates, with error bars representing 95% CIs. **d,** A Schematic of the retinal layers segmented from OCT scans and their names listed from the inner to the outer retina: the Retinal Nerve Fiber Layer (RNFL), Ganglion Cell Layer (GCL), Inner Plexiform Layer (IPL), Inner Nuclear Layer (INL), the complex from the Outer Plexiform Layer to the External Limiting Membrane (OPL to ELM), ELM to Ellipsoid Zone (ELM to EZ), EZ to Retinal Pigment Epithelium (EZ to RPE), and the Retinal Pigment Epithelium + Bruch’s Membrane complex (RPE + Bruchs). **e,** Retinal Layer thickness analysis. The heatmap visualizes the coefficients from eight linear regression models that tested the association between diabetes severity and the thickness of a specific retinal layer, highlighting layer-specific changes in different severity groups. Because CIs cannot be directly visualized within the heatmap, asterisks indicate significant coefficients (adjusted P < 0.05) after Bonferroni correction. **f,** Conditional Association Analysis. Among significant associations identified in the retinal layer thickness analysis, we evaluated the association between diabetes severity and vision outcomes after conditioning on layer thickness, across four vision measures. x-axis shows attenuation of the diabetes-vision association after accounting for retinal thickness, with percentages showing the proportion of the total association accounted for. Error bars represent 95% CIs. The y-axis lists retinal layer-vision outcome pairs with Bonferroni-significant total associations and significant attenuation after conditioning.

In NHANES, survey-weighted group comparisons using the Rao–Scott chi-square test showed a significant association between visual impairment categories and diabetes prevalence (*P* ≤ 2.91 × 10^−2^). The weighted prevalence of diabetes was highest among individuals with irreversible vision impairment (20.31%, 95% CI: 15.85, 24.79), compared with those with correctable impairment (13.32%, 95% CI: 10.99,15.66) or no impairment (14.64%, 95% CI: 12.59, 16.68) (Fig. 4b).

To further characterize this pattern, we utilized the AI-READI dataset, which provides more visual function metrics and granular diabetes severity annotations. Linear regression revealed lower visual function across vision measures with increasing diabetes severity (Fig. 4c). Both insulin-treated and oral medication–treated groups showed significantly reduced photopic contrast sensitivity (log; *n* = 2200) (insulin: β = −0.064, 95% CI: −0.085, −0.044, adjusted *P* ≤ 2.47 × 10^−9^; oral medication: β = −0.025, 95% CI: −0.039, −0.011, adjusted *P* ≤ 2.32 × 10^−10^) and mesopic contrast sensitivity (log; *n* = 2194) (insulin: β = −0.085, 95% CI: −0.110, −0.060, adjusted *P* ≤ 1.60 × 10^−10^; oral medication: β = −0.023, 95% CI: −0.040, −0.005), adjusted *P* ≤ 4.52 × 10^−2^). In addition to contrast sensitivity measures, both reduced photopic vision and mesopic vision were observed in the insulin group. Among these effects, the largest magnitude reduction was observed for mesopic contrast sensitivity in the insulin-treated group.

Associations between retinal structural thickness and diabetes severity were quantified using optical coherence tomography (OCT) across eight retinal layers (Fig. 4d). Significant thinning was observed in the retinal nerve fiber layer (RNFL), photoreceptor inner segment layer (external limiting membrane to ellipsoid zone; ELM–EZ), and photoreceptor outer segment layer (ellipsoid zone to retinal pigment epithelium; EZ–RPE) in both the oral medication-treated and insulin-treated groups (Fig. 4e). The largest effect was the thinning of the photoreceptor outer segment (EZ–RPE, µm; *n* = 1,889), which was evident in the oral medication group and exacerbated in the insulin-dependent group (insulin: β = −1.58, 95%CI: - 2.01 to −1.15, adjusted *P* ≤ 9.59 × 10^−12^; oral medication: β = −0.52, 95% CI: −0.83 to −0.22, adjusted *P* ≤ 5.35 × 10^−3^).

To investigate the association between retinal structure and functional deficits, we performed a conditional association analysis across diabetes severity and vision metrics. This analysis quantified the association between diabetes severity and vision loss after accounting for retinal layer thickness. While multiple associations were significantly reduced after conditioning on retinal thickness (Fig 4f), the most pronounced finding involved EZ–RPE layer thickness and mesopic contrast sensitivity in the insulin-dependent group relative to healthy participants.

Compared to healthy participants, the insulin-dependent group showed the unadjusted association with mesopic contrast sensitivity of (β = −0.086, 95% CI: −0.113, −0.059, adjusted P ≤ 1 × 10⁻³). Conditioning on EZ-RPE thickness yielded a significant attenuation (β = −0.015, 95% CI: −0.020, −0.010, adjusted P ≤ 1 × 10⁻³), indicating that structural thinning of the EZ-RPE layer statistically accounted for 17.31% of the difference in mesopic contrast sensitivity between insulin-dependent and healthy participants. Although these cross-sectional findings reflect statistical associations rather than causal mechanisms, they suggest that thinning of the photoreceptor complex is a measurable structural correlate of functional impairment in advanced diabetes.

### System level performance and Failure-Mode Assessment

This section evaluates the system-level robustness of LATCH, beginning with computational efficiency and followed by performance under stress-testing conditions. End-to-end runtime and LLM token–based costs were measured across 102 clinical hypotheses spanning reproduction, extension, and new-insight analyses (Extended Data Fig. 7). Runtimes ranged from 0.86 to 15.15 min and costs from $0.02 to $0.40, with a general trend toward higher API costs for longer-running queries.

Robustness was assessed through controlled perturbations designed to probe edge-case behavior across three dimensions: Content Validity, Semantic Variability, and Logical Complexity (Fig. 5a). Each safeguard was evaluated independently, as failure at any stage can compromise overall system performance (Extended Data Fig. 4, Supplementary Table 3). A total of 400 stress-test queries were generated across the three evaluation dimensions.

**Fig 5.**
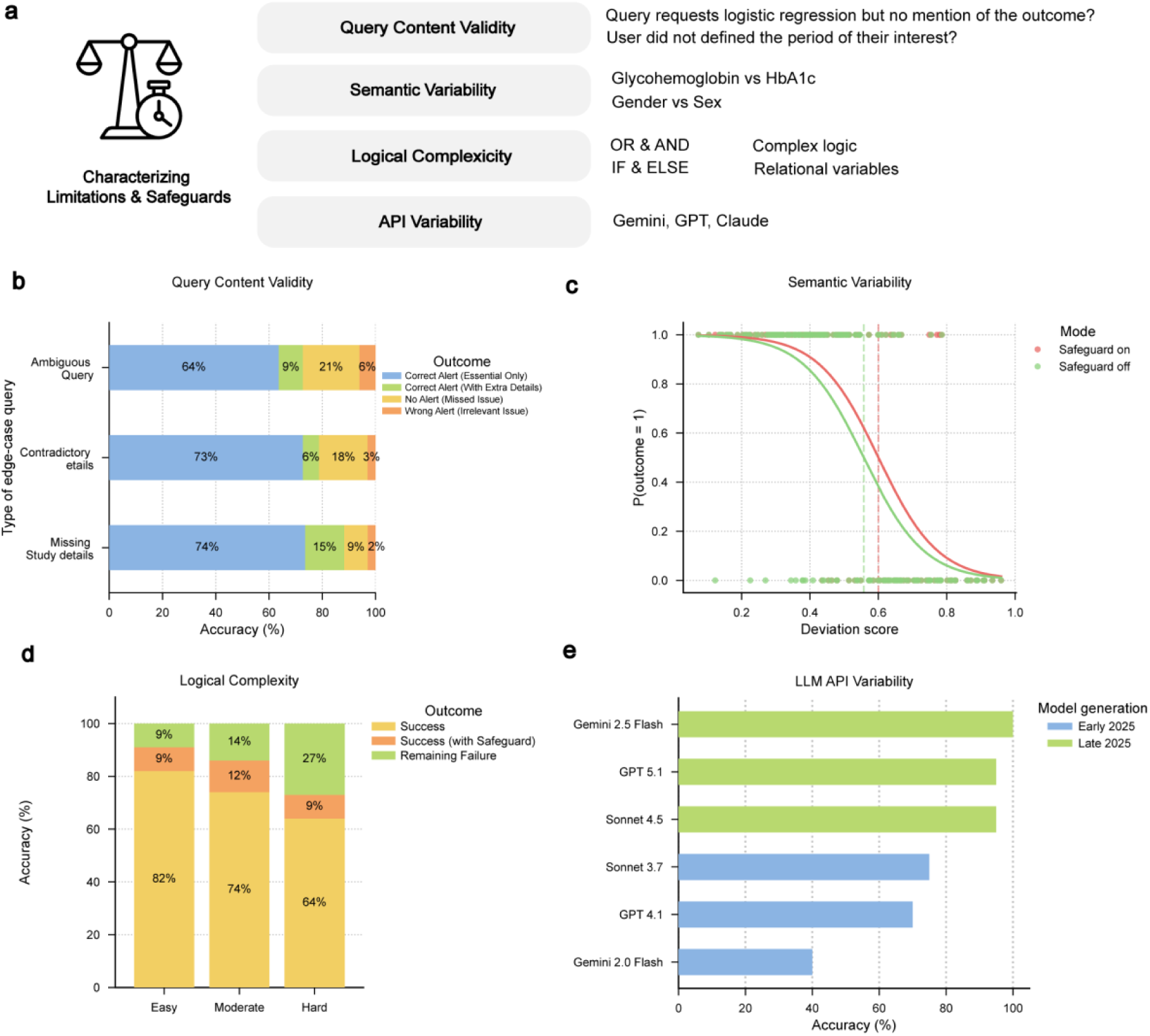
Characterizing Limitations and Safeguards of the LATCH Framework. **a**, Overview of potential sources of failure in translating natural language clinical hypotheses into executable analyses, including query content validity (e.g., missing outcomes or contradictory study details), semantic variability (e.g., substantial variation between clinical terminology and schema variables), logical complexity (e.g., complex logic and relationally derived variables), and API-level variability across model generations. **b**, Query content validity stress test showing outcome distributions for ambiguous queries, contradictory specifications, and missing study details. **c**, Semantic variability analysis illustrating how phrase deviation from schema variables affects outcomes. Sigmoid fits demonstrate that safeguards increase tolerance to semantic deviation, shifting the estimated midpoint parameter (x₀), corresponding to a 50% probability of correct mapping, which is interpreted as the semantic robustness threshold. **d**, Logical complexity evaluation across three difficulty tiers (easy, moderate, hard). Stacked bars indicate first-pass execution success, additional corrections recovered by safeguards, and remaining failures. **e**, API-level variability across contemporary large language models evaluated on identical queries from the reproduction study and prompts highlighting systematic performance differences by model generation.

In Content Validity testing, performance was strongest for detecting missing study details, with an overall accuracy of 89% (74% fully correct and 15% with extraneous information) (Fig. 5b). Performance was lower for ambiguous prompts, with correct vetting responses in 73% of cases (64% correct alerts only and 9% with extraneous information). The system failed to flag issues in 21% of cases and generated irrelevant warnings without the necessary alert in 6% of cases.

Semantic Variability testing evaluated robustness to deviations in phrasing and schema variable naming, with performance expected to decline as semantic deviation increased. For example, replacing “age in years at screening” with a low-deviation variant such as “age at screening” yielded a deviation score of 0.07, whereas a high-deviation substitution such as “time one has been around” produced a score of 0.95. Safeguard interventions improved robustness by shifting the 50% correctness threshold from 0.56 to 0.60 (Fig. 5c). Remaining failures were primarily associated with high-distance substitutions, domain-specific abbreviations, and cases involving multiple closely related variables.

Logical Complexity testing categorized prompts into three tiers: Easy (simple filtering or arithmetic operations), Moderate (sequential dependencies and group-based reference computations), and Difficult (nested negation and hierarchical classification). First-pass execution success rates were 82%, 74%, and 64%, respectively, increasing to 91%, 86%, and 73% after safeguard-based corrections (Fig. 5d). Safeguards improved robustness by correcting syntactic and structural errors, although nested and double-negation queries remain challenging.

To assess API-level variability, reproduction analyses from 20 published studies were repeated using LLMs from different providers and model generations. Two versions per provider, released in early and late 2025, were evaluated. The original reproductions were conducted using Gemini 2.5 Flash, which served as the reference backend during system development, and therefore near-perfect accuracy was expected for this model. Late-2025 models (GPT 5.1 and Claude 4.5 Sonnet) achieved approximately 95% correctness, followed by early-2025 models, including Claude 3.7 Sonnet (75%), GPT 4.1 (70%), and Gemini 2.0 Flash (40%) (Fig. 5e). Earlier-generation models exhibited lower performance than newer models under identical prompts and execution environments.

## DISCUSSION

This study demonstrates that an LLM-assisted framework can accelerate the testing of clinical hypotheses from large health datasets. We show that LATCH can be used to reproduce and extend established analyses and generate new insights through natural language, transforming workflows that typically require time-intensive manual coding into verifiable analyses executed in minutes. Together, these findings suggest a practical step toward more efficient, accessible data-driven medical research by reducing technical and resource barriers to hypothesis testing.

Beyond efficiency, this framework enforces a higher standard of transparency and reproducibility. Reproduction of 20 published analyses exposed methodological ambiguity, where implementation details were often under-specified. Encoding this information within a LATCH prompt and its Analytic Report yields transparent, computationally reproducible analyses.

However, the successful deployment of LATCH in this study was contingent on the use of standardized cross sectional tabular datasets like NHANES and AI-READI. We recognize that this represents an idealized scenario compared to the complexity of “real-world” EHRs, which are notoriously heterogeneous^51,52^. Applying this framework to such environments is a significant challenge and a critical direction for future work as the present findings may overestimate performance in operational clinical environments. Yet this limitation also highlights an opportunity. Our findings suggest a substantial return on investment from data standardization efforts such as the Observational Medical Outcomes Partnership Common Data Model^53^. While standardizing data requires a significant one-time effort, it enables the scalability of evidence with tools like LATCH. Moreover, the incorporation of structured outputs derived from non-tabular sources, such as imaging-derived features transformed into tabular representations, as demonstrated with retinal thickness analysis, suggests a path toward extending LATCH to multimodal data.

An additional limitation is the inherent ambiguity of natural language as it may produce various valid interpretations, highlighting the critical role of human oversight in ensuring alignment with researcher intent. Another limitation stems from the stochastic nature of LLM inference. Although LATCH produces consistent outputs under well-specified prompts, model responses remain probabilistic, and commercially hosted LLMs may undergo undisclosed architectural or inference-level updates that alter behavior over time. These may limit strict reproducibility and highlight the importance of version tracking and record keeping.

A related consideration is the potential for rapid, automated analyses, which increase the risk of false-positive findings and may lower barriers to uncontrolled exploratory testing. Although LATCH provides an auditable record of analytic steps, it does not eliminate risks associated with multiple testing or selective reporting, underscoring the need for prespecification of outcomes and adherence to statistical principles. These risks can be mitigated by requiring external validation of findings in independent datasets, preregistration, and transparent tracking of hypotheses through registries that record all attempted analyses and promote appropriate correction for multiple testing. As LLM-based analytic tools continue to evolve, such challenges are likely to become unavoidable, and proactively addressing them through deliberate methodological standards will be essential.

As an LLM-assisted framework that manages the end-to-end pipeline from a natural language question to a verifiable statistical result, LATCH offers a practical vision for the future of biomedical research. It shifts scientific effort from repetitive coding toward hypothesis formulation, analysis evaluation, and result interpretation, emphasizing researcher focus on insight generation, review, and validation rather than manual implementation. Reproducibility becomes an intrinsic property of the analytic workflow rather than a retrospective concern. Together, these advances point toward a more efficient, transparent, and scalable ecosystem for translating clinical data into actionable knowledge, with the potential to accelerate scientific discovery and its impact on human health.

## METHODS

### Dataset

#### Overview

Prior to analysis, all datasets undergo a one-time pre-processing procedure designed to generate a comprehensive metadata artifact: the Schema Summary Table. This table enables the Large Language Model (LLM) to interpret the dataset’s structure without any exposure to the underlying patient-level data. The procedure utilizes the dataset’s publicly available data dictionary and documentation and consists of two primary stages (Extended Data Fig. 1).

First, in the code-to-text translation stage, the data dictionary is programmatically parsed to map technical variable identifiers (e.g., DIQ010) and their corresponding value encodings (e.g., 1) to their full natural language descriptions (“Doctor told you have diabetes”) and labels (“Yes”). Second, during the Schema Summary Table construction, metadata information is then consolidated and structured into a single reference table where columns include variable names, table names, natural language descriptions, data types, and example values, as available.

The resulting Schema Summary Table serves as the sole source of information for the LLM during the subsequent query interpretation phase, thereby ensuring a strict separation between the model’s semantic understanding of the data schema and the confidential patient data itself. All processing was conducted in Python using the Pandas^54^ library.

#### NHANES

The data acquisition process involved the programmatic retrieval of all available XPT (SAS transport) files from the National Health and Nutrition Examination Survey (NHANES) public data portal spanning 1999–2023, a total of 12 survey cycles across five major categories: Demographics, Dietary, Examination, Laboratory, and Questionnaire, which yielded 1,562 tables covering 128,809 unique participants. Variable metadata and codebooks were obtained directly from the official NHANES documentation pages. For each dataset, corresponding codebook pages were scraped to extract definitions, Statistical Analysis System (SAS) labels, English-language descriptions, and value encodings. Parts of the NHANES ingestion pipeline were adapted by translating selected functions from the nhanesA^55^ R package into Python. SAS natural language variable labels were selected as SQL column names because they were both semantically descriptive and compatible with database’s identifier length constraints. A minority of variables with identical codes but inconsistent SAS labels across years were resolved using a frequency-based harmonization strategy, where the most common label was adopted. This resulted in a single, unified metadata file that serves as the basis for the Schema Summary Table used by LATCH.

#### NHANES Mortality data

To enrich the dataset for survival analyses, associated mortality data were obtained from the NHANES Linked Mortality Files which are available for survey cycles 1999–2018 and had 101,316 unique participants. Mortality status, cause-of-death codes, and follow-up time were extracted from .dat files, which were converted into tables in csv files and were linked to NHANES data via the respondent sequence number (SEQN). Finally, all tables in csv files, including survey data and mortality files, were loaded into a database.

#### AI-READI Clinical data

The clinical data for the AI-READI Year 3 cohort^23^ were obtained following approval through the standard AI-READI data access application process. The cohort consisted of 2,280 participants. The dataset was provided in the Observational Medical Outcomes Partnership (OMOP) Common Data Model (CDM) format and included the person, measurement, observation, and condition_occurrence tables. The procedure_occurrence table was not included in the final analytic dataset due to the presence of only one recorded procedure across the cohort, resulting in negligible informational value for feature generation. To create a unified, analysis-ready table from these files, a multi-step processing workflow was executed. All column names and variable labels were standardized to lowercase and replaced special characters with underscores, ensuring consistency for data joins and transformations. Concurrently, data quality was ensured by first removing duplicate rows based on the combination of person_id, description, and value, and then by excluding irrelevant or ambiguous survey items (e.g., timestamps, vague frequency questions) based on a predefined list. Observational records were filtered and merged with the external OMOP concept dictionary to retrieve standardized human-readable labels associated with OMOP Concept IDs. A publicly available OMOP documentation table^56^ was used to construct a concept mapping dictionary. To accommodate nested REDCap-style value dictionaries, the mapping procedure converted raw numeric and string-based encodings into standardized categorical representations, such as mapping binary indicators (for example, “1”) to interpretable labels (“Yes”), thereby preserving semantic consistency across variables. Variables with excessively long descriptive labels were shortened to comply with character-length limitations imposed by downstream reporting and analysis platforms. Additionally, variables with unclear or noninformative labels were renamed to improve interpretability. A structural transformation involved reshaping the dataset from the native OMOP long format, in which each row represents a single observation, into a wide-format representation. The observation and measurement tables were independently pivoted using person_id as the primary index, with standardized clinical variable labels serving as column headers.

#### AI-READI OCT Imaging Data

The retinal imaging data for the AI-READI Year 3 cohort^23,57^ were obtained following approval through the standard AI-READI data access application process. Macular 6 × 6 mm structural optical coherence tomography (OCT) volumes were acquired using the Topcon Maestro2 imaging platform. For each scan, two corresponding Digital Imaging and Communications in Medicine (DICOM) files^58^ were provided: a structural OCT volume and a segmentation (SEG) file containing multiple retinal boundary surfaces. In total, 4,506 paired OCT–SEG DICOM file sets were included in the dataset. File correspondence between OCT and SEG data was established using a study manifest table that included the relative paths to each associated pair. Segmentation and OCT volumes were read using pydicom^59^, and only successfully paired files from a predefined subject list were analyzed. The segmentation pixel data were extracted as a 3-dimensional array with dimensions corresponding to retinal boundaries, B-scans, and A-scans.

Nine retinal layer boundary labels were obtained from the DICOM metadata from the tag Segment Sequence. For each adjacent boundary pair, layer thickness (in pixels) was computed at every A-scan and B-scan position as the absolute difference in depth between boundary coordinates. The resulting per-scan dataset included thickness measurements for all retinal layers in native pixel units. Physical scaling information was extracted directly from the OCT DICOM metadata using the PixelMeasuresSequence tag. Axial spacing, lateral spacing, and slice thickness were used to convert the derived pixel thicknesses into micrometers. For eyes with multiple scans, thickness maps were averaged at the eye level; subsequently, averages across both eyes were computed when possible, to obtain per-person imaging features. The final dataset contains standardized retinal layer thickness values in micrometers, organized consistently by subject ID and eye laterality, and is suitable for downstream statistical modeling and multimodal integration.

### LATCH

LATCH is an automated framework designed to streamline the analysis of tabular health data through a standardized and transparent pipeline. A central design philosophy of the system is the selective use of LLMs. The system employs LLMs exclusively for tasks requiring semantic reasoning, such as interpreting ambiguous clinical language, while utilizing deterministic, rule-based systems for statistical execution and recording the process. This hybrid approach allows LATCH to maintain the flexibility of natural language understanding while adhering to the strict reproducibility standards required for medical research.

#### Architecture

LATCH comprises five specialized modules that mirror the standard scientific workflow: Planner, Variable Mapper, Data Engine, Statistics Engine and Reporter (Fig. 1a, Extended Data Fig. 2). These modules are organized into two functional layers. The first consists of three hybrid semantic modules (Planner, Variable Mapper and Data Engine), which integrate LLM-based semantic reasoning with rule-based constraint enforcement to generate executable code and logic from natural language input. The second layer comprises two fully deterministic execution modules (Statistics Engine and Reporter), which perform statistical computation, aggregation and reporting using fixed, reproducible pipelines.

A central Python orchestration layer coordinates this workflow, integrating heterogeneous software components, such as LLM APIs, PostgreSQL^60^, and R, via libraries such as SQLAlchemy^61^ and rpy2. Crucially, this architecture enforces data privacy by design. The LLM is used only to generate code and logic, and it never accesses the patient database directly. Data processing occurs locally within the secure environment, ensuring that patient-level information is never exposed to external API calls.

All components within the LLM-assisted semantic layer (Planner, Variable Mapper, and Data Engine) were driven by a structured prompt template that combined step-by-step task instructions with a one-shot example^62^ demonstrating the expected input-output format (Supplementary Fig. 1,2,3). This standardized prompting strategy improved output consistency, reduced formatting and logical errors, and enforced structured, machine-readable responses.

#### Planner

The Planner module uses a large language model (Google Gemini 2.5 Flash; temperature = 0) in two sequential processing stages. In the first stage, free-text study hypotheses are standardized into a structured text with a field-based analytical specification. This process extracts key analytical components, including the selected dataset, study period, analysis method, and method-specific variables (for example, predictors, covariates, and outcomes), as required by the analysis type. During this step, the system’s safeguard identifies missing or inconsistent study specifications, flags unsupported analytical methods, and prompts users for clarification when necessary. Subsequently, the structured text is converted into a machine-readable JSON study plan. This dual-stage design improves JSON generation reliability and ensures that the study plan is aligned with the requirements of the selected analytical model.

#### Variable Mapper

The Variable Mapper module connects clinical concepts in the study plan to database variables. To prevent hallucinations, this module uses a hybrid retrieval process rather than direct LLM generation. First, keywords are extracted from the JSON study plan in a rule-based manner. For each keyword, a semantic search model (all-MiniLM-L6-v2^63^) retrieves candidate variables from the schema summary based on cosine similarity. Because the schema summary contains a large number of variables across the full dataset, this distance-based filtering step is used to identify the top 40 candidate variables and to ensure compatibility with LLM context window constraints. Second, an LLM performs a classification to select the most contextually appropriate variable from this set of candidates. Each candidate variable is presented to the LLM as a filtered subset of the schema summary table, with available metadata fields such as table name, variable name, example values, and descriptions, allowing the model to compare candidates and select the best-aligned variable. This process is performed for each keyword in the study plan and produces a validated mapping between clinical concepts (e.g., “fasting blood sugar level”) in the user query and database variable (e.g., fasting_glucose_mmol_l), ensuring that downstream analyses are grounded in the available schema. These mappings are incorporated into the JSON study plan in a rule-based manner, resulting in an enriched specification in which each keyword is paired to its corresponding table and column identifiers.

#### Data Engine

The Data Engine module ingests the enriched JSON study plan to programmatically construct the analytic cohort. It begins with a rule-based step that aggregates mapped variables across data cycles and produces a master table. Performing this step outside the LLM ensures structural integrity, reduces context window overhead, and reserves LLM compute for higher-level reasoning, as the procedure involves only deterministic variable aggregation. Subsequently, the schema of the master table is provided to the LLM, which functions as a logic synthesizer and generates SQL queries to implement study-specific inclusion/exclusion criteria, perform feature engineering, and format variables according to the required statistical analysis method. The generated SQL query is executed locally via SQLAlchemy, and results are stored as a pandas DataFrame.

#### Statistics Engine

The Statistics Engine module executes the statistical analysis on the prepared pandas^54^ DataFrame outputted by the SQL query. LATCH has the separation of the flexible, semantic query interpretation that involve LLM-assisted steps from rigid statistical execution. This design ensures consistency and reproducibility by eliminating analytic variability, such as inconsistent statistical packages, parameters, or significance thresholds, which would invalidate comparisons across separate analyses.

The module processes the input data through a sequential, automated analytical pipeline. On the pandas DataFrame outputted by the SQL query, feature classification occurs to label variables as numerical, binary, or categorical. If needed, reference levels for categorical predictors are automatically selected based on clinically meaningful keywords (e.g., “healthy”, “none”, “control”, “Q1”) or, when absent, the most frequent level. Statistical analyses are executed according to the analysis method specified during the Planner phase, which was selected from a predefined set of available statistical analyses. Supported analyses include logistic regression, linear regression, Cox proportional hazards, prevalence estimation, group-wise numerical comparisons, stratified logistic regression, and mediation analysis.

For unweighted analyses, the module utilizes lm for linear regression, glm (binomial family) for logistic regression, and coxph from R’s survival^64^ package for time-to-event modeling. Group-based comparisons of continuous variables are conducted using Welch’s t-test for two-group comparisons and one-way Analysis of Variance (ANOVA) for multi-group scenarios. Categorical associations are evaluated using Pearson’s chi-square test. Mediation analyses are performed using the mediate package. All confidence intervals are calculated at the 95% level, and effect estimates are reported as odds ratios (OR) or hazard ratios (HR) where applicable. For complex survey designs (e.g., NHANES), the module utilizes the R survey^65^ package to account for multi-stage sampling. Regression analyses are executed via svyglm and svycoxph. Weighted prevalence estimates and group-specific summary statistics are computed using svymean and svyby. Between-group differences are evaluated using design-based hypothesis tests, including svyttest for two-group comparisons and svychisq for categorical group comparisons. Survey weighting is enforced through fixed, deterministic templates applied as a rule-based layer on top of the standard query generation pipeline, conforming to NHANES analytic guidelines and automatically filtering participants without valid sampling weights while ensuring inclusion of all required survey design variables prior to statistical execution.

In case of missing data, for survey-weighted analyses, the engine defaults to complete-case analysis to maintain the integrity of the survey design structure, which can be compromised under standard imputation. For unweighted analyses, complete-case analysis is the default, with optional support for Multiple Imputation via Chained Equations (MICE)^66^ or simple imputation (mean/mode) for exploratory sensitivity analyses.

#### Reporter

The final module generates a comprehensive Analytic Report that serves as an immutable audit trail of the full analytical workflow. This component operates as a rule-based logging system that records all intermediate and final pipeline artifacts in a structured comma-separated values (CSV) format. Each column in the report corresponds to a predefined metadata field, and all inputs and outputs are captured without modification. The report aggregates structured outputs from each stage of the pipeline, including the parsed user hypothesis, cohort definition parameters, executed SQL queries, selected schema variables, and final statistical results such as model coefficients and summary tables. Each field is written to predefined columns using fixed formatting rules, ensuring that all inputs and outputs remain directly traceable, verifiable, and reproducible.

#### Safeguard System

A safeguard mechanism is integrated across the LLM-assisted components of LATCH, specifically the Planner, Variable Mapper, and Data Engine, to enhance operational robustness.. The safeguards (Extended Data Fig. 4) are applied at three LLM-assisted stages of the workflow, as described below:

Safeguard 1. At the Planner step, this performs user input vetting, evaluating the user’s free-text request against high-level study design requirements (e.g., compatibility between analysis method and outcome type) and verifying dataset and study-year availability. Requests that are ambiguous, incomplete, or unsupported trigger a clarification prompt with specific guidance before execution proceeds.

Safeguard 2. During the Variable Mapping step, this applies a rule-based filter that first identifies candidate variables that are consistently defined across the specified study period, mitigating errors due to variable drift across data cycles. When multiple consistent candidates exist, an LLM selects the most contextually appropriate variable based on the full study specification; when no single consistent variable is available, year-specific mappings are applied and logged for transparency.

Safeguard 3. In the Data Engine module, this performs iterative SQL Self-Correction in the Data Engine module. If a generated SQL query fails execution, the database error message and schema context are fed back to the LLM to refine the query. This automated correction loop iterates for up to three attempts, resolving syntax or logic errors before terminating.

### Reproduction of Published Studies

#### Literature Search and Curation of Reproduction Studies

To evaluate the reproducibility of NHANES-based diabetes research, we curated a representative cohort of 20 studies (Extended Data Fig. 5). PubMed was searched for articles published between 2010 and 2025 using combinations of the following terms: (“National Health and Nutrition Examination Survey” OR “NHANES”) AND “diabetes”, together with outcome-specific keywords for diabetic retinopathy (“diabetic retinopathy”), diabetic kidney disease (“diabetic kidney disease”), all-cause mortality(“all-cause mortality”), and mental or cognitive health (“depression” OR “cognitive” OR “mental” OR “cognition”). Search terms restricted to title and abstract fields. Inclusion was limited to studies with full-text availability to ensure the presence of sufficient methodological detail for reproduction.

Because the objective was to assess reproduction feasibility rather than provide a comprehensive meta-analysis, we employed a ranked-sampling approach. The retrieved articles per topic were ranked by Google Scholar citation count in descending order. We screened these results sequentially and applied the following exclusion criteria: use of non-standard or restricted NHANES data or external data sources; populations not restricted to individuals with diabetes; redundancy in exposure-outcome relationships already represented by a higher-ranked study; focus on outcomes outside the four domains of interest; methodological descriptions insufficient for exact reproduction, including ambiguous definitions of inclusion/exclusion criteria, exposures, outcomes, or covariates; involved data structures unsuitable for standardized multi-cycle analysis, such as highly granular dietary or prescription drug records; and statistical methods unsupported by the LATCH framework.

Following this ranked screening procedure, the first five eligible studies were selected for each of the diabetic retinopathy, diabetic kidney disease, and mental or cognitive health outcome categories. To demonstrate time-to-event analyses (e.g., Cox regression), three all-cause mortality studies were included. In addition, two supplementary diabetic retinopathy studies were incorporated because they contained overlapping risk factor definitions shared between the NHANES and AI-READI datasets, enabling planned cross-dataset validation experiments. In total, 20 studies were selected for reproduction.

#### Running Reproduction Analysis on LATCH

The primary objective of this reproduction study was to establish a standardized benchmark for the LATCH system’s analytical fidelity. We began by systematically decomposing the methodologies of the original publications into a structured framework comprising cohort definitions, inclusion/exclusion criteria, predictor and outcome variables (if applicable), and variable transformations. For all derived variables, we explicitly defined the computational logic and underlying raw data components within the prompt to ensure the LATCH system accurately reconstructed the original study’s metrics. These parameters were synthesized into natural language prompts (Supplementary Table 1) to guide the LATCH system’s automated execution, ensuring that each analysis remained strictly aligned with the original study periods and datasets.

While the original publications typically reported results across three levels of complexity, univariate, demographic-adjusted, and fully adjusted models, we focused our primary reproduction on the demographic-adjusted models to ensure a high degree of analytical comparability. By prioritizing demographic models over “fully adjusted” specifications, which often involve heterogeneous covariate definitions and under-reported missing data handling protocols, we minimized the risk of misattributing discrepancies to irreproducibility when they may have stemmed from unstated modeling choices. We standardized these models using a consistent set of variables, specifically “Gender,” “Age at screening,” and “Race/Hispanic Origin,” and ensured all laboratory variables included units. This standardized approach provided a uniform baseline for validation. For the two studies ^31,38^ where a demographic-only model was unavailable, we utilized the most conservative (least-adjusted) specification reported to ensure a fair assessment of the original findings. For studies utilizing complex survey designs, we specified survey weights within the prompt as described in the original publications. In cases of methodological ambiguity, we prioritized a conservative default (e.g., examination weights if exam values were used as a variable) to ensure the analytic sample size remained consistent with the source material and to prevent artificial sample loss. The reproduction was performed by inputting these individual prompts, one per analysis, directly into the LATCH system; the subsequent data retrieval, processing, statistical analysis, and results generation were automated.

#### Concordance Evaluation

Concordance between results generated by LATCH and those reported in the original publications was evaluated using multiple complementary metrics designed to capture agreement in direction, statistical significance, effect magnitude, and cohort construction.

Primary concordance measures were applied primarily to logistic regression and Cox proportional hazards models, which constituted the majority of reproduced analyses and provided standardized relative effect estimates suitable for direct comparison. These measures included assessment of effect direction, determining whether both analyses indicated increased or decreased risk; statistical significance concordance, evaluating whether both results were significant (*p* < 0.05) or both were non-significant; and overlap of 95% confidence intervals when available, focusing on the most extreme exposure exposure category (for example, the highest quartile or tertile). Additional descriptive measures were used to support interpretation, including comparison of analytic sample sizes between LATCH and the published studies to assess alignment in cohort construction, and calculation of relative effect size ratios (published estimate divided by the LATCH estimate) when effect measures were directly comparable^44^.

When discordance was observed, a root-cause analysis was conducted by manually reviewing both the constructed prompts and the original publications to identify potential sources of discrepancy, such as ambiguous cohort definitions, unclear variable transformations, or insufficiently specified data processing procedures.

Because outcome reporting formats varied substantially across studies, concordance criteria were adapted accordingly (Supplementary Table 2). For prevalence or percentage-based outcomes lacking confidence intervals, agreement was assessed using absolute differences, with values considered concordant if the difference was within 1 percentage point. For group comparisons reported only as means and standard deviations, concordance focused on agreement in direction of between-group difference and confidence interval overlap when available. For linear regression analyses reporting beta coefficients with confidence intervals and *p-*values, concordance was evaluated based on effect direction, confidence interval overlap, and statistical significance, while relative effect size ratios were applied only when effect scales were directly comparable.

After applying the concordance criteria specific to each analysis type, a replication outcome was assigned for each reproduced analysis. An analysis was classified as fully concordant if all applicable replication assessment criteria were satisfied. If one or more criteria were met but at least one criterion failed, the result was classified as partially concordant. Analyses failing all primary concordance criteria were classified as non-concordant.

#### Repetition Study

To assess output consistency and sources of variability, we executed each of the reproduction analysis from 20 studies independently three times using LATCH under identical input conditions. Reproduction outcomes were classified as consistent when all runs produced equivalent cohort definitions and statistical results, and as variable when discrepancies were observed across runs. For variable cases, discrepancies were manually reviewed and categorized into three sources: Valid interpretability that reflects reasonable alternative implementations of analytic details not explicitly specified in the original publication; Underspecified methods where insufficient methodological detail precluded a unique reconstruction; Misimplementation which corresponds to incorrect application of clearly defined analytic rules.

### Extension of Existing Studies

The framework’s utility for accelerating scientific inquiry was demonstrated by extending the findings through modifications to the original natural language queries. These extensions were designed to probe the generalizability and robustness of the original associations by evaluating common limitations such as dataset specificity, temporal change, patient heterogeneity, and reliance on less granular outcomes. Specifically, the extension study comprised four distinct analyses, moving beyond the scope of the original publications: cross-dataset generalizability for testing external validity across independent cohorts; temporal consistency for assessing effect consistency across different time periods; stratified analysis with confounder adjustment; and granularity enhancement for providing higher-resolution outcome definitions.

#### Cross-Dataset Generalizability

To assess external validity, we evaluated previously reported risk factors for diabetic retinopathy across two independent cohorts: NHANES and AI-READI. This analysis included three risk factors from the reproduction study^26,29,30^ (originally derived using NHANES data) and ten additional risk factors identified from a published American Academy of Ophthalmology (AAO)’s Diabetic Retinopathy Preferred Practice Pattern^45^. To enable a same-setting comparison, NHANES analyses were replicated within the AI-READI dataset by re-initializing the LATCH reproduction workflow with modified natural language prompts. Adjustment for demographic covariates was restricted to age, as additional demographic variables such as sex and race were not available in the publicly accessible AI-READI dataset. These prompts aligned study periods with cohort-specific data availability and defined the AI-READI “age” variable as the direct equivalent to the NHANES age variable. To facilitate a direct comparison of effect sizes, the prompts specified unweighted logistic regression models for both datasets, as the survey design variables required for weighting were not applicable for the AI-READI cohort. Participants who had missing values in our risk factor of interest, diabetic retinopathy status or without diabetes were excluded. Most variables of interest, including BMI, hemoglobin, serum albumin, kidney disease status, glycohemoglobin, blood pressure, insulin use, LDL cholesterol, smoking history, and diabetic retinopathy status, were directly transferable across datasets through LATCH’s automated variable mapping without requiring dataset-specific redefinition. However, to maintain analytical integrity, the system’s automated safeguards trigger a warning when multiple potential variable matches are detected across data cycles. This occurred specifically for physical activity, where the presence of multiple overlapping NHANES questionnaire variables prompted a supervised refinement of the natural language instruction. We explicitly specified “Physical activity (vigorous activity questionnaire)” rather than a generic “Physical activity” prompt for the NHANES analyses. This human-in-the-loop review ensured consistent variable alignment across datasets. A post hoc Bonferroni correction was applied to adjust for multiple hypothesis testing across 13 risk factors.

#### Temporal Consistency

To assess the stability of risk factor associations over time, we selected studies on all-cause mortality in diabetic patients with extended observation periods: one investigating the Prognostic Nutritional Index^46^ and another evaluating the Weight-Adjusted Waist Index^47^. Using weighted Cox proportional hazards regression with mortality as the primary outcome, we adjusted for demographic variables across an aligned, overlapping study window from 2005 to 2018. To evaluate temporal consistency, we employed a two-tiered analytical approach consisting of a pooled analysis and a cycle-specific analysis. In the pooled analysis, a unified model was executed across the entire combined study period to establish a baseline effect size. This was complemented by a cycle-specific analysis, where the LATCH workflow was initialized to perform independent analyses for each of the seven biennial NHANES cycles within that window (2005-2006, 2007-2008, 2009-2010, 2011-2012, 2013-2014, 2015-2016, and 2017-2018). This allowed us to investigate potential fluctuations in these associations and determine whether the identified risk factors remained consistent across successive cohorts or were subject to temporal variation. The only difference between input prompts were the study periods in the natural language input. The resulting hazard ratios and 95% confidence intervals for each cycle, comparing the highest exposure category (Q4) with the lowest (Q1), were plotted in a line graph where the Y-axis represented hazard ratio and the X-axis represented the NHANES cycle to visually assess temporal trends or consistency in effect sizes.

#### Stratified Analysis with Confounder Adjustment

Although serum albumin is known to be biologically associated with renal function^49,50^, the initial study on the relationship between albumin and diabetic retinopathy did not account for kidney disease status.^26^ To provide additional adjustment for potential confounding and to investigate patient heterogeneity in this association, we extended the original analytical framework by adding a chronic kidney disease (CKD) status as a covariate and modifying the model from standard logistic regression to stratified logistic regression. CKD was defined as an estimated glomerular filtration rate (eGFR) < 60 mL/min/1.73 m² or a urine albumin-to-creatinine ratio (UACR) ≥ 30 mg/g using relevant lab values. Attenuation of the serum albumin effect and changes in statistical significance were evaluated to assess confounding by kidney disease. Comparisons of effect estimates across strata were used to determine whether the association differed by underlying kidney dysfunction.

#### Granularity Enhancement

To provide a more detailed, stage-specific clinical characterization, we refined the analysis by moving beyond binary outcome definitions. Whereas the original reproduction study^29^ categorized diabetic retinopathy (DR) as a binary outcome, we leveraged the available granular DR severity categories, including no DR, mild, moderate/severe DR, and proliferative DR. This approach allowed us to investigate the systematic trend of different variables, including hemoglobin, duration of diabetes, glycated hemoglobin (HbA1c), red cell distribution width (RDW), across the increasing severity stages of DR. These relationships were visually presented in plots where mean lab values were plotted against the ordered DR severity levels, providing a more detailed, stage-specific profile than dichotomous comparisons.

### Generation of New Insights

We used LATCH to generate new insights through a sequential, hypothesis-driven workflow. Analyses were performed using natural language specifications, enabling automated end-to-end execution while maintaining analytical transparency. We first examined diabetes prevalence across vision impairment groups in NHANES and then extended these analyses to the AI-READI cohort, which provides detailed ophthalmic phenotyping and retinal structural measures. All steps, including data selection, variable definitions, exclusion criteria, and statistical modeling, were defined via natural language prompts.

#### Trend Identification (NHANES)

For population-level trend identification, we used NHANES data from five survey cycles (1999-2008) which reported vision data. We included participants aged 20 years or older with complete data for both presenting and post-refraction visual acuity in both eyes, and we excluded individuals missing diabetes related information, defined as absence of both glycohemoglobin (HbA1c) measurements and self-reported physician diagnosis of diabetes. Diabetes was a binary variable, defined as HbA1c ≥ 6.5% or clinician-reported diabetes. Visual status was derived using the better-seeing eye and categorized into a three-level group variable: no visual impairment (20/40 or better), correctable visual impairment (presenting acuity worse than 20/40 but post-refraction acuity 20/40 or better), and nonrefractive visual impairment (worse than 20/40 even after refraction). Using NHANES survey weights, exam weights, and accounting for the complex sampling design, we estimated weighted prevalence of diabetes across visual impairment groups and survey cycles to assess population-level trends in vision-associated diabetes burden.

#### Visual Deficit Analysis (AI-READI)

To investigate functional visual deficits associated with diabetes more precisely, we performed linear regression analyses using the AI-READI dataset collected during 2023-2025. Participants were excluded if data were missing for the diabetes severity group or for required visual outcome and covariate variables. Three visual outcomes were analyzed. Photopic vision was defined using photopic logMAR acuity as the better (lower) value of the two eyes, multiplied by −1 so that higher scores indicate better vision. Mesopic vision was defined analogously to day vision, using mesopic logMAR values with the lower value multiplied by −1. Contrast sensitivity was defined as the higher value between the two eyes using log contrast sensitivity measurements. Both photopic and mesopic contrast sensitivity data was used. In all visual function models, the primary predictor was study group, representing diabetes severity, and age and ocular comorbidities (reported diagnosis of glaucoma, cataract, diabetic retinopathy, age-related macular degeneration, and retinal vascular occlusion) were included as covariates. A post hoc Bonferroni correction was applied to adjust for multiple hypothesis testing across 4 vision metrics.

#### Structural Retinal Layer Thickness Analysis (AI-READI)

To characterize structural and abnormalities in diabetes, we analyzed retinal layer thicknesses in micrometers extracted from OCT scans, including retinal nerve fiber layer (RNFL), ganglion cell layer (GCL), inner plexiform layer (IPL), inner nuclear layer (INL), outer plexiform layer to external limiting membrane (OPL-ELM), photoreceptor inner segment layer (external limiting membrane to ellipsoid zone; ELM-EZ), photoreceptor outer segment layer (ellipsoid zone to retinal pigment epithelium; EZ-RPE), and retinal pigment epithelium layer (RPE+Bruch’s). Participants were excluded if data were missing for the diabetes severity group or for required retinal layer information and covariate variables. For each layer, we fit a separate linear regression model with study group as the main predictor while adjusting for age and ocular comorbidities (reported diagnosis of glaucoma, cataract, diabetic retinopathy, age-related macular degeneration, and retinal vascular occlusion). This allowed us to identify layers that are selectively affected in diabetes, providing structural correlates to the observed functional deficits. A post hoc Bonferroni correction was applied to adjust for multiple hypothesis testing across 8 retinal layers.

#### Conditional Association analysis (AI-READI)

Among significant associations identified in the structural retinal layer thickness analysis (RNFL, ELM-EZ, EZ-RPE, and RPE+Bruch’s), we conducted a mediation-style conditional association analysis using the LATCH mediation module to evaluate whether retinal layer thickness statistically accounted for the association between diabetes severity and vision outcomes. Given the cross-sectional nature of the data and lack of temporal ordering, results were interpreted as conditional associations rather than causal mediation effects. Participants with complete data on diabetes severity (X), retinal layer thickness (M), vision outcomes (Y), and age and ocular covariates were included. Using ordinary least squares (OLS) regression in R, we fit sequential models and estimated indirect and direct association components using the *mediate* function from the mediation R package, as implemented within the LATCH mediation module. Sequential models were specified as follows:

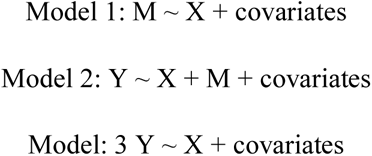

Regression coefficients were summarized as path a (association between X and M from Model 1, path b (association between M and Y from Model 2, and c′ (conditional association between X and Y from Model 2). The total association (c) was obtained from Model 3. The indirect association was quantified as the product a × b, representing the attenuation of the total diabetes-vision association when accounting for retinal layer thickness. This attenuation was reported as an absolute effect size (β), expressed in the original outcome scale (for example, log contrast sensitivity units). In addition, the proportion of the total association between diabetes severity and vision outcomes explained by retinal thickness was calculated as (a × b) / c and reported as a percentage. Confidence intervals for the indirect association (a × b) were estimated using bootstrap resampling, implemented through the mediate function. A post hoc Bonferroni correction was applied to adjust for multiple hypothesis testing across 16 analyses derived from four retinal layers and four vision metrics. For visualization, we plotted only retinal layer-vision pairs in which both the total diabetes severity-vision association and the attenuation after conditioning on retinal thickness were statistically significant after correction, and where the direction of the diabetes effect was consistent with reduced visual function.

### System Stress Test and Safeguard Assessment

To evaluate the operational boundaries of LATCH and its robustness under edge-case conditions, we conducted a comprehensive stress-testing and safeguard assessment. System robustness was examined through controlled perturbations targeting three LLM-integrated modules: the Planner, the Variable Mapper, and the Data Engine, together with their associated safeguard mechanisms (Extended Data Fig 4). Each module was evaluated independently, as failures at any stage can propagate downstream and compromise overall system performance. The evaluation was anchored on a set of validated queries derived from the three clinical domains included in the reproduction study: diabetic retinopathy, diabetic kidney disease, and depression. From the query set, one representative query per domain was selected as a baseline for systematic perturbation. Although these baseline queries had previously demonstrated structural completeness, logical coherence, and strong alignment with the underlying database schema, they were intentionally modified to generate 400 challenging variants designed to stress different system components. Content validity testing focused on the Planner safeguard module (n = 100), evaluating the system’s ability to detect missing, contradictory, or ambiguous analytical specifications from the user. Semantic variability testing targeted the Variable Mapper safeguard (n = 200), introducing diverse natural language reformulations to assess tolerance to schema misalignment and linguistic variation. Logical complexity testing focused on the Data Engine safeguard (n = 100), probing the system’s capacity to preserve logical consistency and correctly translate complex analytical intent into executable SQL queries. By systematically perturbing the factual, linguistic, and logical structure of validated inputs, this evaluation framework enabled characterization of LATCH’s operational limits under potentially challenging conditions.

#### Query Content Validity

This evaluation assessed safeguard that performs user input vetting at the Planner stage. A total of 100 queries were systematically perturbed by introducing missing critical study elements (e.g., removal of the study period or undefined outcomes), ambiguous phrasing (e.g., “exclude sick people”), or contradictory specifications (e.g., requesting linear regression for a binary outcome). Safeguard responses were classified into four outcome categories: Correct Alert (Essential Only), in which the safeguard correctly identified the issue and provided only the necessary corrective guidance; Correct Alert (With Extra Detail), in which the safeguard correctly identified the issue and included unnecessary information; No Alert (Missed Issue), in which the safeguard failed to detect the problem and proceeded with the flawed query without warning; and Wrong Alert (Irrelevant issue), in which the safeguard failed to identify the true issue and instead generated irrelevant or unnecessary feedback. Results were summarized as the percentage of queries falling into each outcome category.

#### Semantic Variability

This evaluation examined the robustness of the automated schema selector to natural language variation and quantified the contribution of the Variable Mapper safeguard. A systematic perturbation study was conducted in which 200 variant prompts were generated from variable phrases appearing in the validated queries from the reproduction study. Each Variant Prompt was created by applying a single-word or single-phrase substitution within the original query template using a curated synonym list for relevant variables (for example, replacing “serum albumin” with “blood albumin level”).

Each Variant Prompt was processed by the Variable Mapper with the safeguard enabled, and outputs were evaluated using a binary correctness metric (1 = correct mapping, 0 = incorrect mapping), defined by whether the perturbed phrase was successfully mapped to the appropriate underlying database variable. Outputs that required safeguard intervention to achieve correct mapping were labeled Safeguard Required, whereas outputs that achieved correct mapping without intervention were labeled Safeguard Not Required.

The magnitude of semantic perturbation was quantified using a deviation score ranging from 0 to 1. Sentence embeddings were generated for the original schema phrase (A) and the perturbed variant phrase (B) using the Sentence-BERT (SBERT) framework with the all-mpnet-base-v2 model^67^. The deviation score was computed as the inverse cosine similarity:

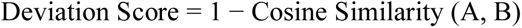

To isolate the effect of the internal robustness mechanism, the dataset was segmented into two groups: Safeguard Required (correctness achieved only after safeguard intervention) and Safeguard Not Required (correctness achieved without intervention). For each group, the relationship between semantic deviation (x) and the probability of successful mapping (P(Correct) = y) was modeled using a sigmoid function fitted via nonlinear least-squares optimization (scipy.optimize.curve_fit). The estimated midpoint parameter (x₀), corresponding to a 50% probability of correct mapping, was interpreted as the semantic robustness threshold. This framework enabled quantitative comparison of how the schema matching safeguard shifted system performance under increasing semantic perturbation. Results were visualized by plotting raw binary correctness outcomes alongside the fitted sigmoid curves for each group.

#### Logical Complexity

This evaluation assessed the safeguard in the Data Engine step by examining the robustness of the LLM-based text-to-SQL generation module and the effectiveness of the iterative SQL correction loop under increasing logical complexity. A total of 100 queries were systematically perturbed to isolate the impact of logical structure by selectively modifying cohort selection logic and variable engineering operations. Three levels of logical complexity were defined. Easy difficulty included simple filtering and calculations. Moderate difficulty required logics that involved sequential dependencies and group-based reference computations. High difficulty involved the implementation of nested negation and hierarchical classification. System performance was measured using the final SQL execution success rate, defined as whether the generated query both executed without error and correctly implemented the intended cohort selection and variable engineering logic. Safeguard efficacy was quantified as the proportion of initially failed SQL queries that were successfully repaired within the three-attempt limit of the iterative correction loop.

#### API Variability

This evaluation characterized LATCH system behavior under substitution of the core large language model across all functional pipeline stages. Using the same set queries used for the 20 reproduction studies, we evaluated system performance across multiple contemporary LLM backends spanning different providers and model generations in addition to Gemini 2.5 Flash which was originally used for the reproduction study. For each provider, two representative model versions released in early and late 2025 were selected to capture generational differences in capability. The evaluated models included Claude 4.5 Sonnet (2025-09-29), Claude 3.7 Sonnet (2025-02-19), Gemini 2.0 Flash (February 2025), GPT-4.1 (2025-04-14), and GPT-5.1 (2025-11-13).

The original reproduction pipeline was developed and validated using Gemini 2.5 Flash, which served as the reference backend for the system and was therefore treated as the baseline implementation. For cross-model evaluation, the original queries corresponding to each of the 20 studies were executed without any perturbation using each alternative LLM backend. Identical prompt templates and inputs were used across all pipeline modules, including the Planner, Variable Mapper, and Data Engine, to ensure that observed differences were attributable solely to backend model substitution. Resulting system outputs were compared for correctness and reproducibility. All safeguard mechanisms were enabled during this evaluation to preserve standard operational conditions. The primary objective was to quantify end-to-end system sensitivity to backend model substitution rather than to optimize prompt performance or independently benchmark individual safeguards.

Performance was assessed using task-level correctness, defined as successful reproduction of the intended analytical logic and study outputs under identical execution conditions. To account for minor implementation-level variability that does not alter analytical meaning or cohort definition, a limited set of semantically equivalent outputs was accepted as correct. Specifically, four cases were classified as valid despite non-identical formulations. These included: (1) use of mathematically equivalent cohort threshold expressions (for example, < versus <= when boundary values were not present in the data); (2) minor decimal precision differences in cutoff values (for example, 0.33 versus 0.3333); (3) substitution of closely related clinical variables that reflected the same clinical construct and did not alter the intended clinical interpretation; (4) logically equivalent pregnancy exclusion logic, such as explicitly excluding pregnant participants versus including only non-pregnant participants. These variations were considered analytically equivalent because they produced functionally equivalent cohort definitions and downstream results under the study data distribution. This experiment design enabled controlled comparison of system robustness across LLM providers and model generations.

## Data availability

The NHANES dataset and its metadata are publicly available at https://wwwn.cdc.gov/nchs/nhanes/. NHANES mortality data are publicly available at https://ftp.cdc.gov/pub/Health_Statistics/NCHS/datalinkage/linked_mortality/. The clinical and retinal imaging data for the AI-READI Year 3 cohort (released on November 17, 2025) are available to the public on the FAIR Data Innovations Hub (https://fairhub.io/datasets/3) upon completion of an application process. Access to the dataset was granted following completion of the standard data access and approval procedures. The metadata for clinical data was obtained from the publicly available documentation table at https://docs.aireadi.org/v3-omopAndClinicalTable.

## Code availability

The code used in this study is publicly available at https://github.com/latch-project. The repository includes the LATCH framework source code, data preprocessing code, and the prompts and scripts used for the various analyses presented in the paper, along with their associated outputs.

## Author contributions

N. Gim and A.Y. Lee conceived the study. N. Gim developed and implemented the LATCH system. I. Gim contributed to the technical design and implementation of the LLM-integrated agentic core system architecture and the computational environment. N. Gim conducted experiments using LATCH on clinical research applications. Y. Jiang contributed to the review of the statistical module. A.Y. Lee supervised the work. N. Gim, I. Gim, Y. Jiang, Y. Kihara, Y. Wu, M. Blazes, A.Y. Lee, and C.S. Lee contributed to the writing, revision, and approval of the final manuscript.

---

**Extended Data Fig.1.**
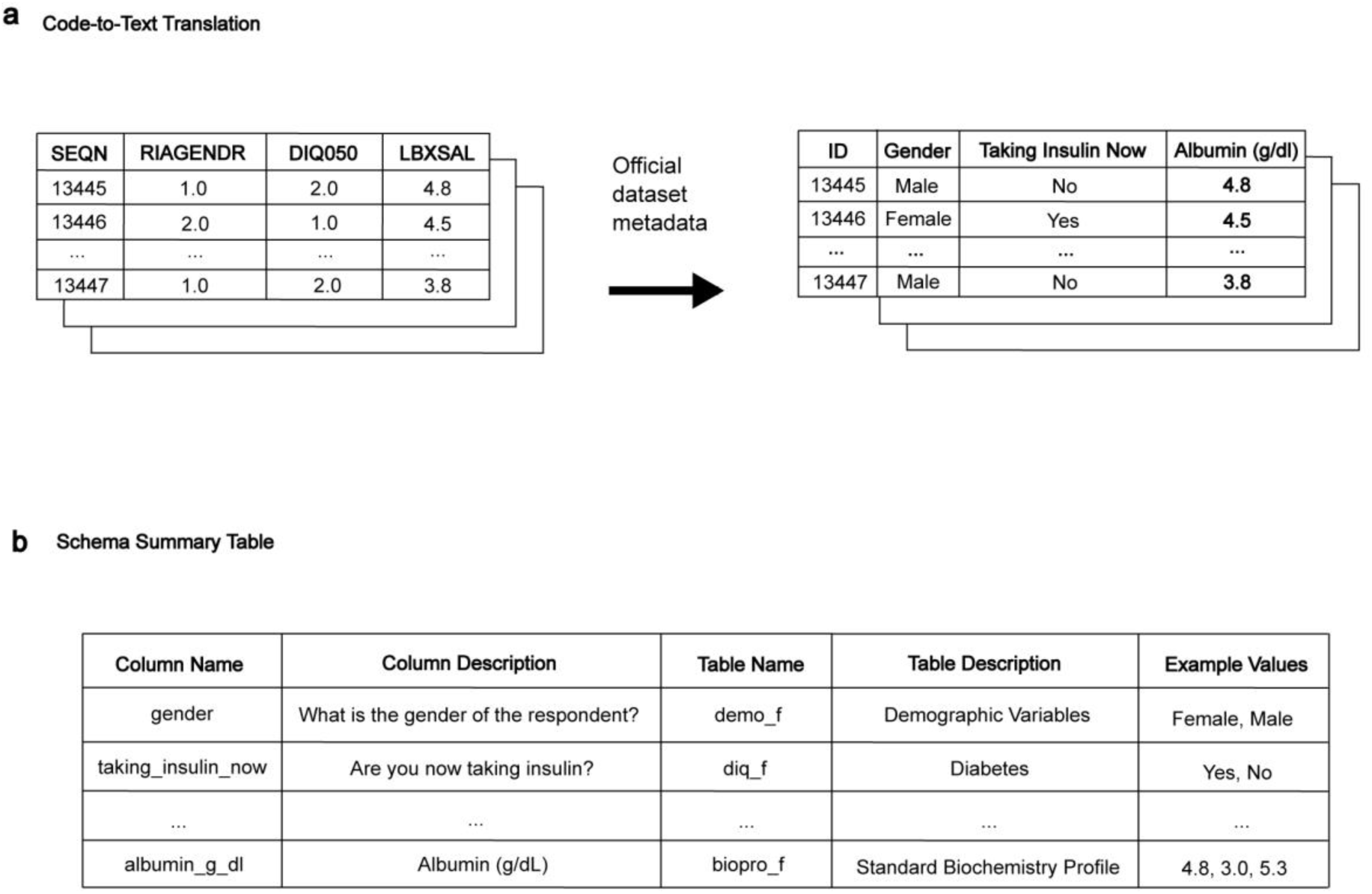
Overview of the data processing workflow. The Code-to-Text Translation step converts coded variable names into natural language descriptions using official metadata typically provided with the dataset. This enables the large language model (LLM) to interpret variable meanings without prior knowledge of the coding system and ensuring that generated analytic code remains interpretable during human review. A Schema Summary Table is then created to summarize available variables and their attributes. During routine operation, only this table, no patient-level data, is shared with the LLM. This schema summary forms the basis for subsequent schema-grounding steps, allowing LATCH to map natural language concepts to corresponding database variables.

**Extended Data Fig.2.**
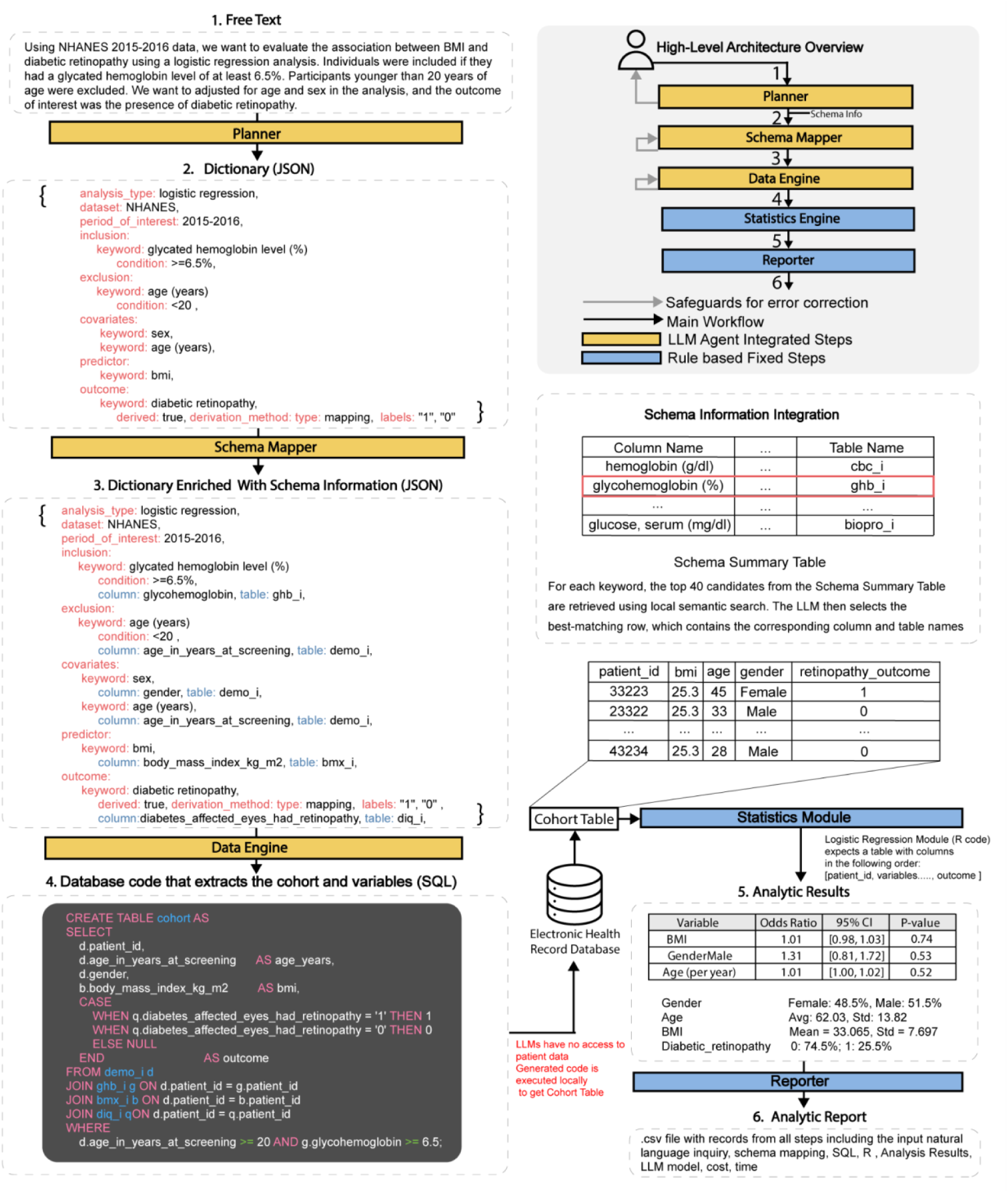
Stepwise LATCH analytic pipeline. (1) A natural language research question is converted into (2) a structured JSON encoding cohort definitions and analytic parameters. This specification is (3) enriched by mapping concepts to dataset-specific schema elements and used to generate (4) executable SQL that extracts the cohort. The resulting table is processed by statistical modules to produce (5) summaries and regression outputs. All workflow stages are documented in (6) an Analytic Report. All code (SQL and R) is executed locally, and LLMs do not access patient-level data.

**Extended Data Fig.3.**
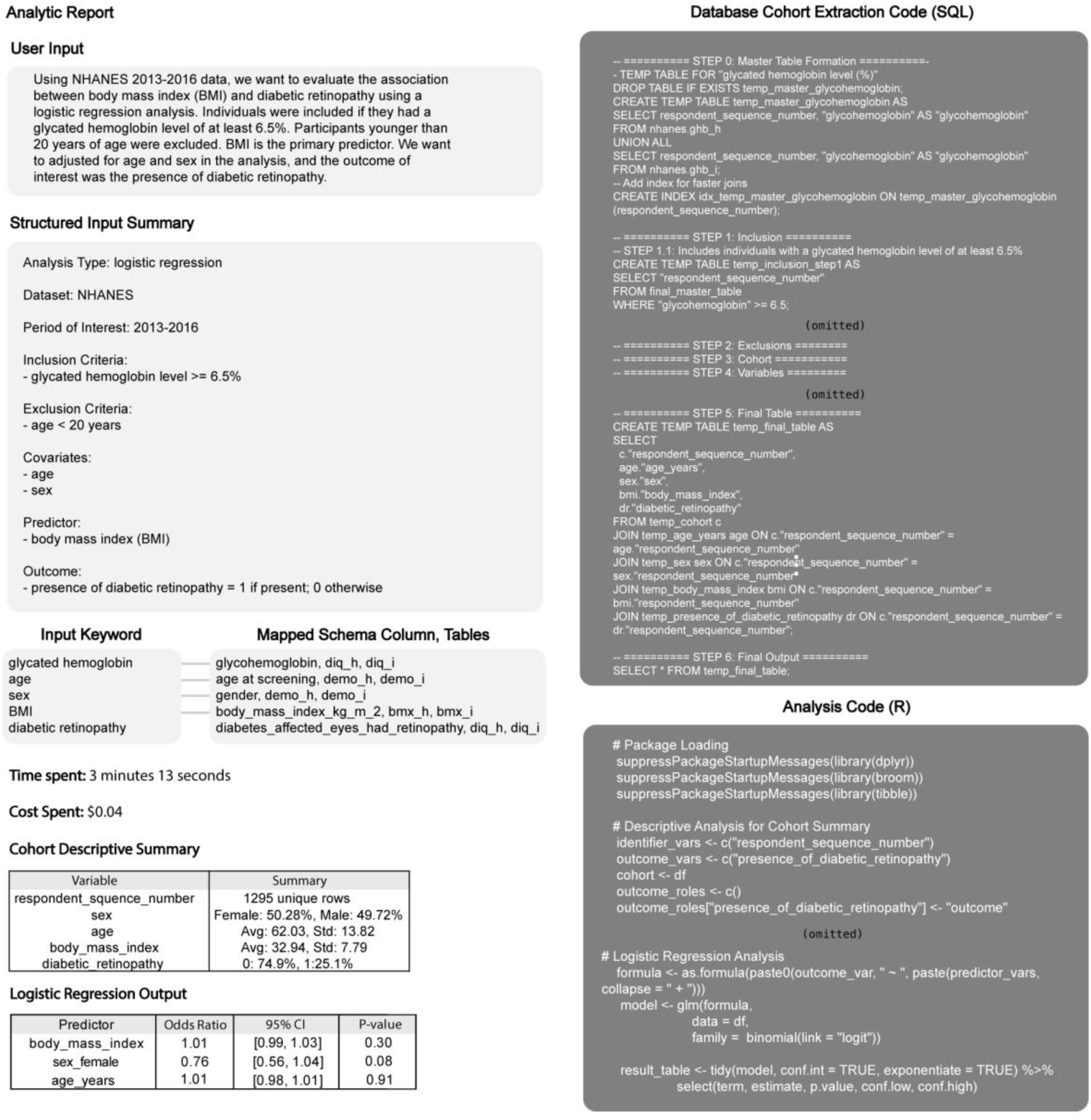
Example Analytic Report generated by LATCH. Illustrative analytic receipt documenting an end-to-end analysis generated from a natural language research query. The receipt includes the original user input, a structured study design summary specifying analysis type, dataset, inclusion and exclusion criteria, covariates, predictor, and outcome, and the corresponding schema mappings matching user-specified concepts to database tables and columns. Generated cohort extraction code (SQL) is shown alongside downstream statistical analysis code (R), enabling transparent reconstruction of cohort definition and modeling steps. Execution time, API cost, cohort characteristics, and regression outputs, are recorded to support auditability, reproducibility, and tracking of each analysis.

**Extended Data Fig.4.**
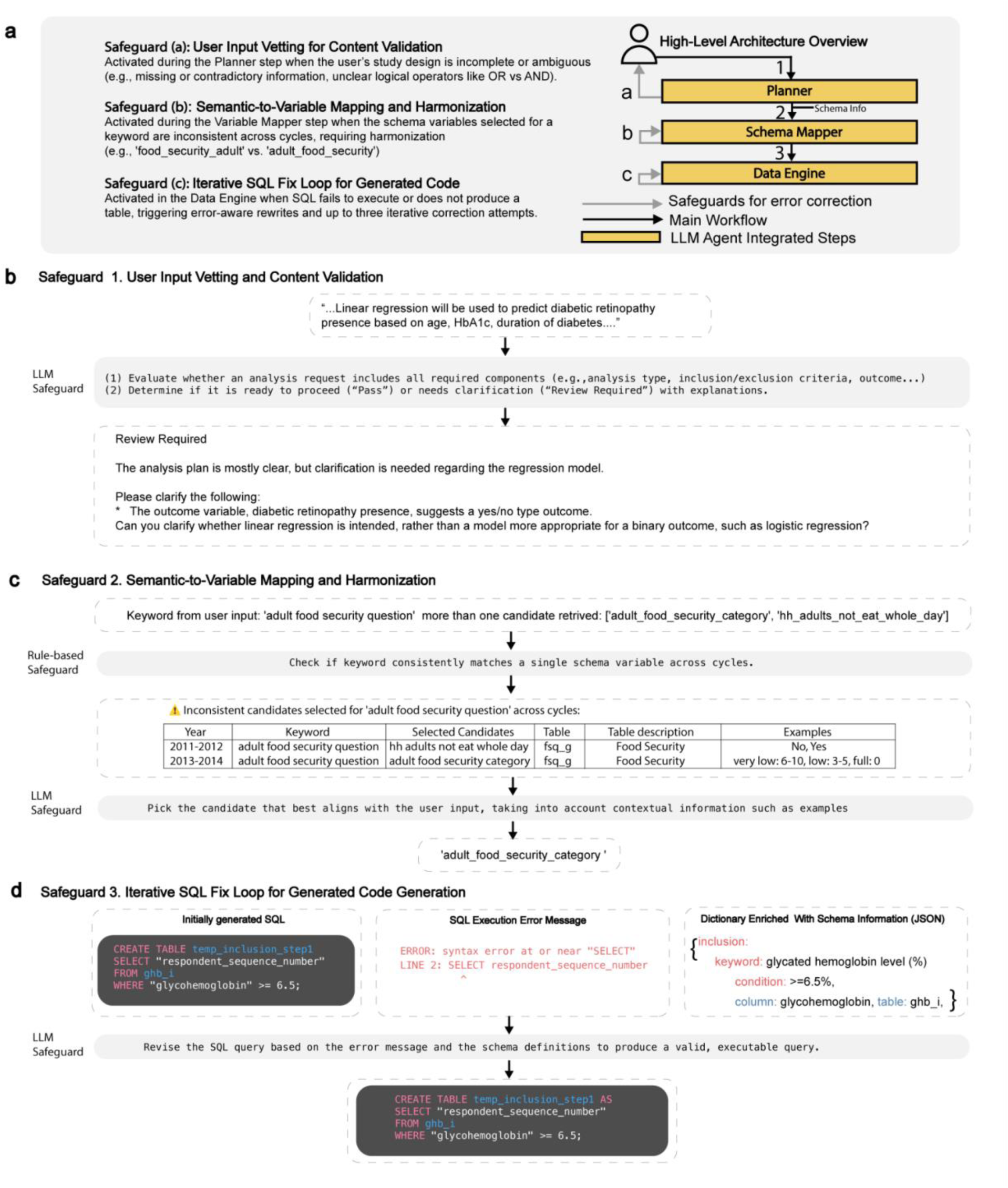
Safeguards in LLM integrated modules in LATCH. **a,** High-level architecture showing the Planner, Schema Mapper, and Data Engine with integrated safeguards for error detection and correction. **b,** Safeguard 1 vets user input and requests clarification when specifications are not sufficient. **c,** Safeguard 2 enforces consistent semantic-to-schema variable mapping across database cycles. **d,** Safeguard 3 iteratively corrects SQL using execution feedback and schema metadata from earlier steps.

**Extended Data Fig.5.**
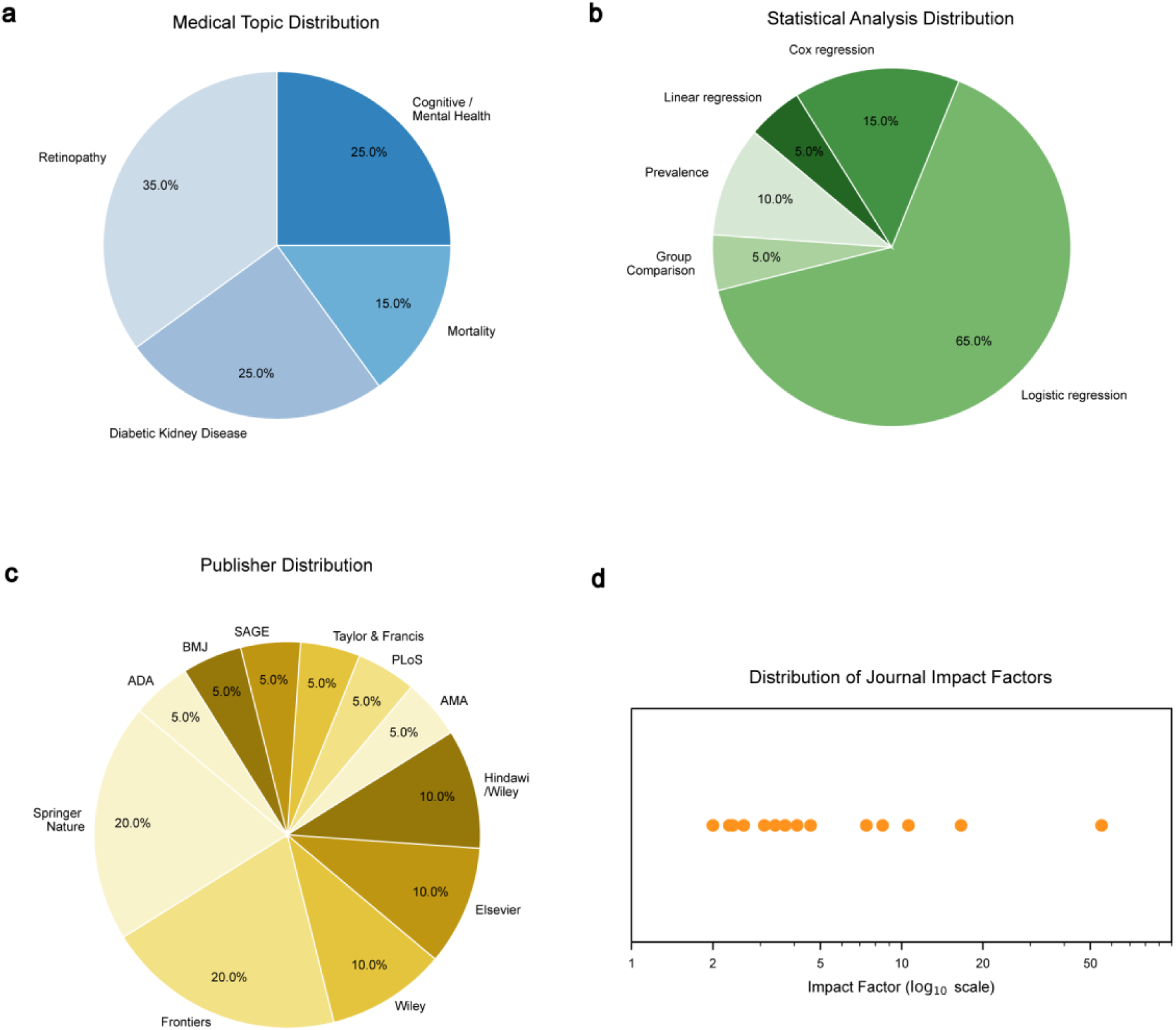
Characteristics of the reproduction studies included in the evaluation. **a,** Medical topic distribution of the 20 selected replication studies, spanning diabetic retinopathy, diabetic kidney disease, cognitive or mental health outcomes, and all-cause mortality. **b,** Statistical analysis distribution of the analyses LATCH reproduced. **c,** Publisher distribution of the reproduction studies, including a range of biomedical journals and publishers. **d,** Distribution of journal impact factors for the reproduction studies, shown on the raw scale with logarithmic spacing.

**Extended Data Fig.6.**
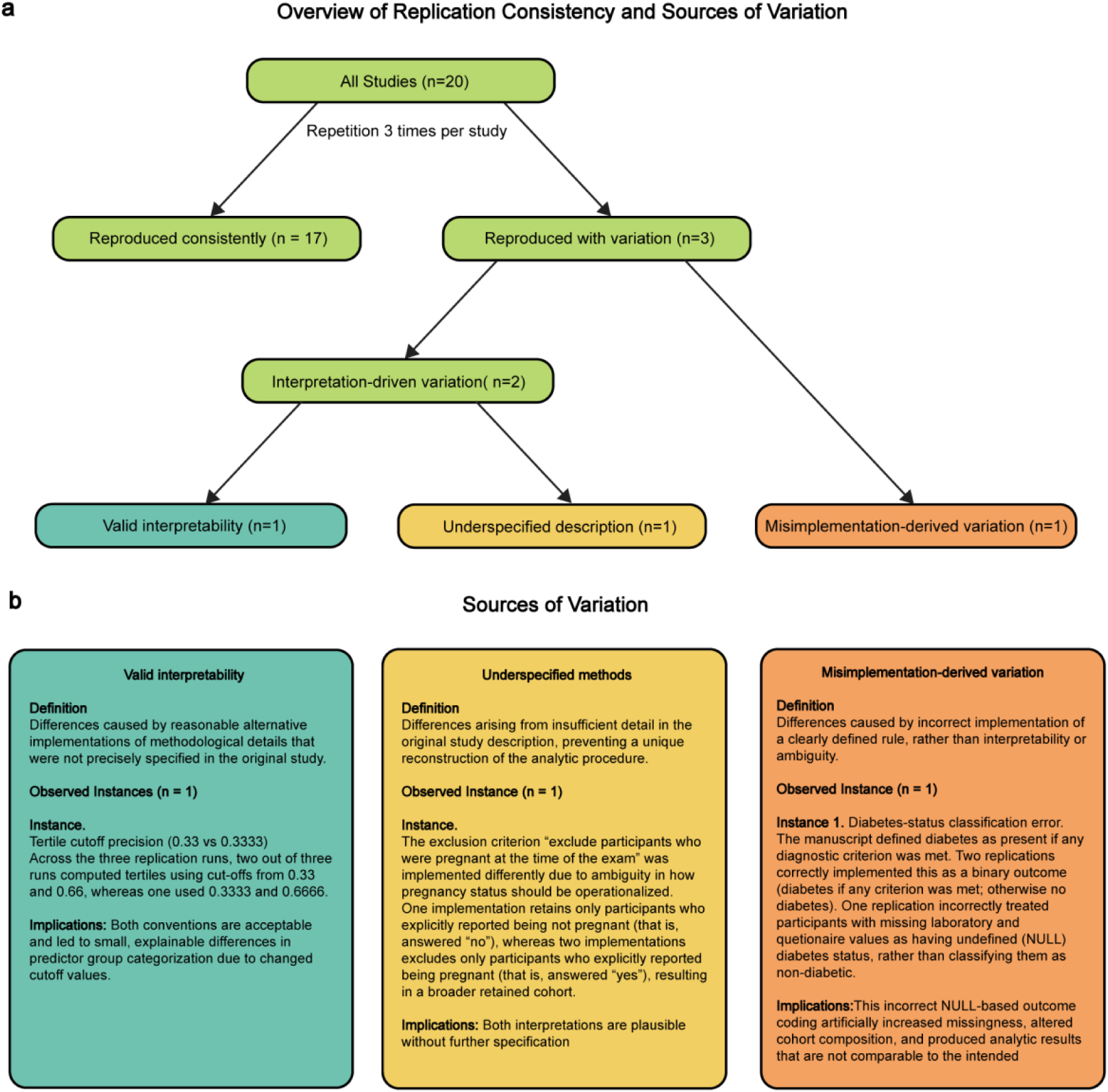
Repetition consistency and sources of analytic variation. **a,** Overview of replication outcomes across 20 studies, each repeated three times to assess consistency given the non-deterministic nature of LLM outputs. Seventeen studies reproduced consistent results, whereas three exhibited variation. Among variable cases, two were attributable to interpretation-driven differences, stemming from valid alternative implementations or underspecified descriptions, and one resulted from a misimplementation-derived error. **b,** Three illustrative examples of analytic variation, one each representing a valid interpretive difference, an underspecified method, and a misimplementation-derived variation, with definitions, observed instances, and implications for analytic reproducibility.

**Extended Data Fig.7.**
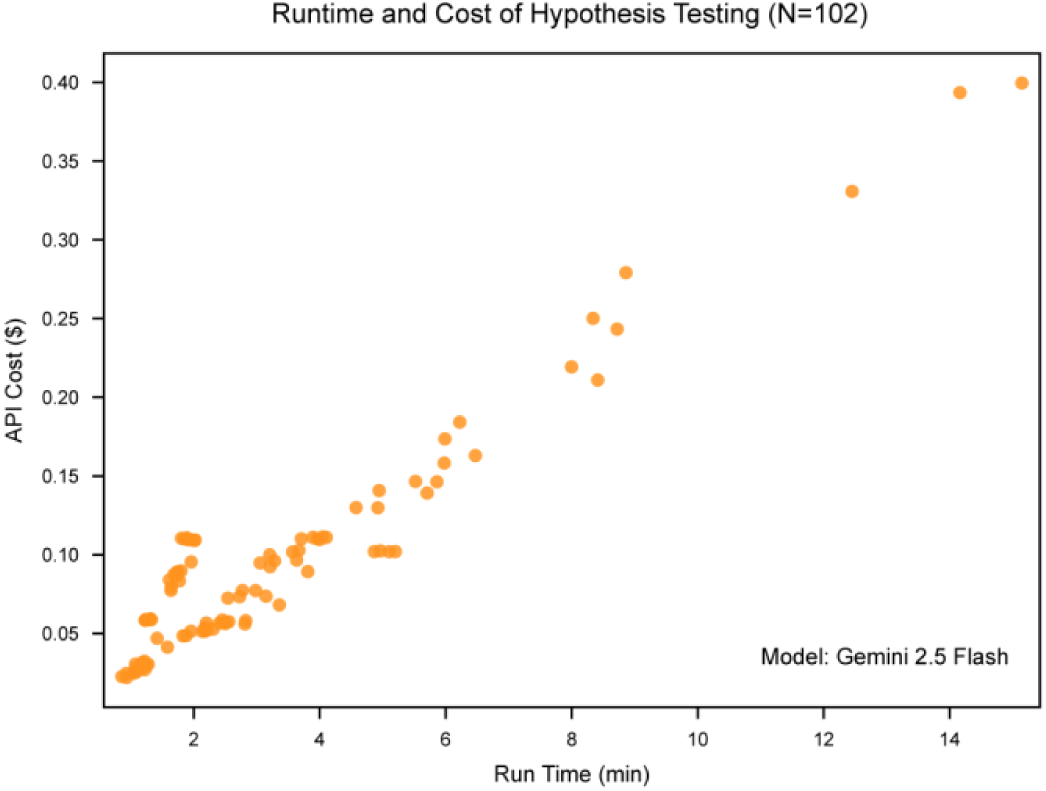
End-to-end runtime and API cost of automated hypothesis testing. Scatter plot showing measured end-to-end runtimes and LLM token-based API costs across 102 clinical hypotheses spanning reproduction, extension, and new-insight analyses. Runtimes ranged from 0.86 to 15.15 min and costs from $0.02 to $0.40. Each point represents a single hypothesis test. Reported costs and time reflect estimated API usage for the Gemini Flash 2.5 model. Reported runtimes and costs are approximate and may vary with network conditions and token usage, which can differ slightly across runs even for identical queries.

**Supplementary Fig.1.**
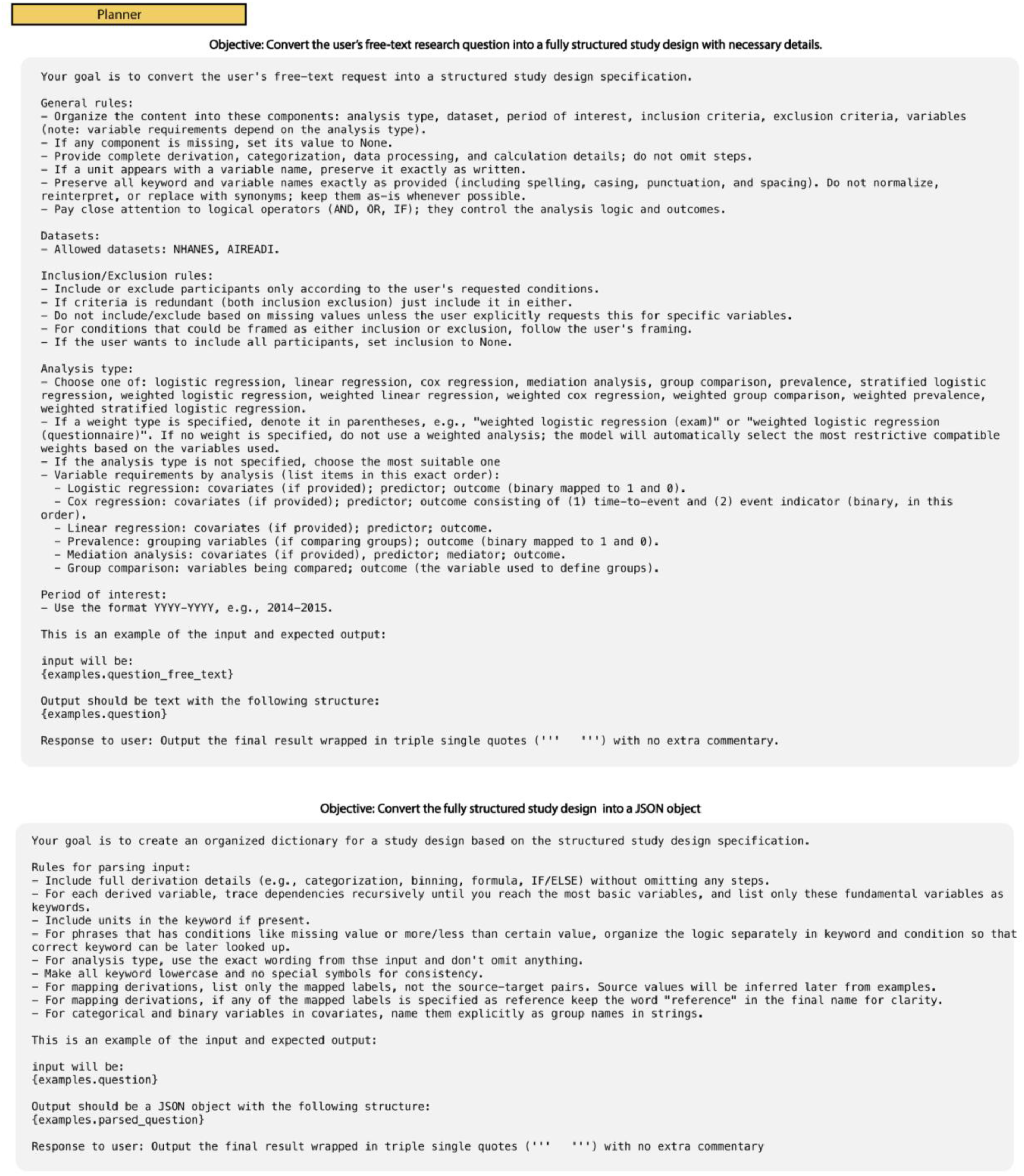
Prompt used in the Planner module. The LLM operates in two stages: first converting user free-text input into structured study design specifications and then transforming the structured output into a JSON object. A one-shot example is provided for each stage. The same prompt was used across all experiments in this study.

**Supplementary Fig.2.**
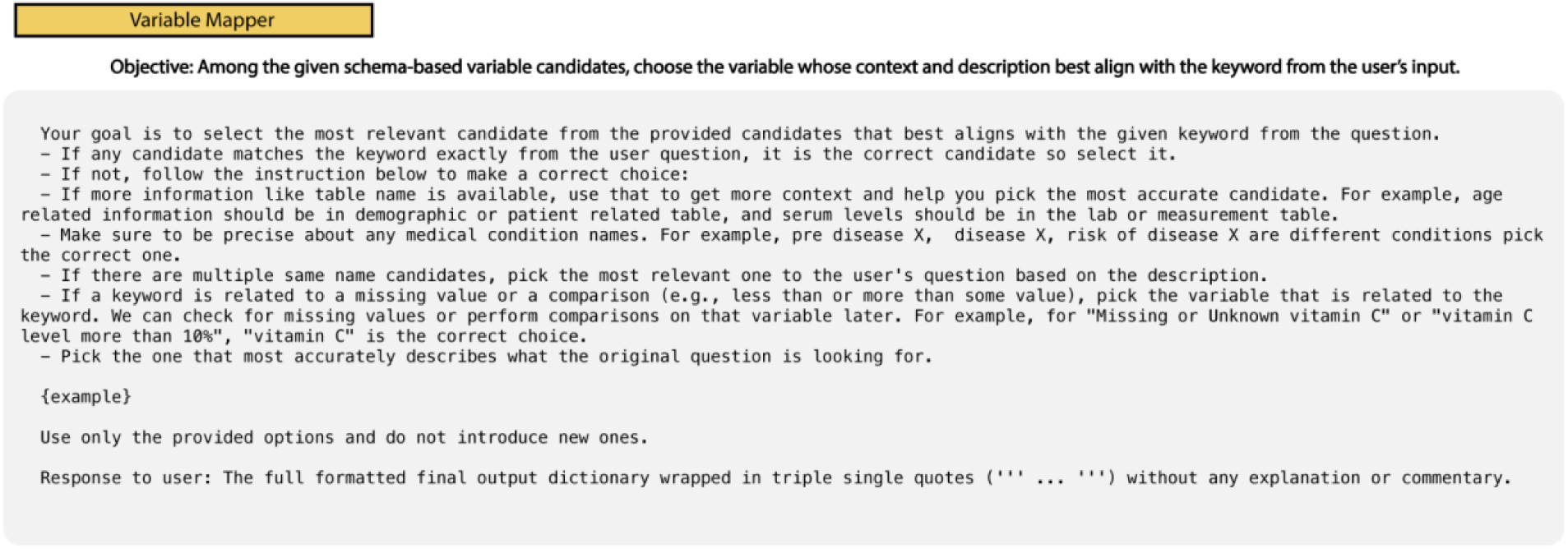
Prompt used in the Variable Mapper module. The LLM selects variables from schema-based candidates by matching the input keyword and original user text. A one-shot example is included. The same prompt was applied across all experiments in this study.

**Supplementary Fig.3.**
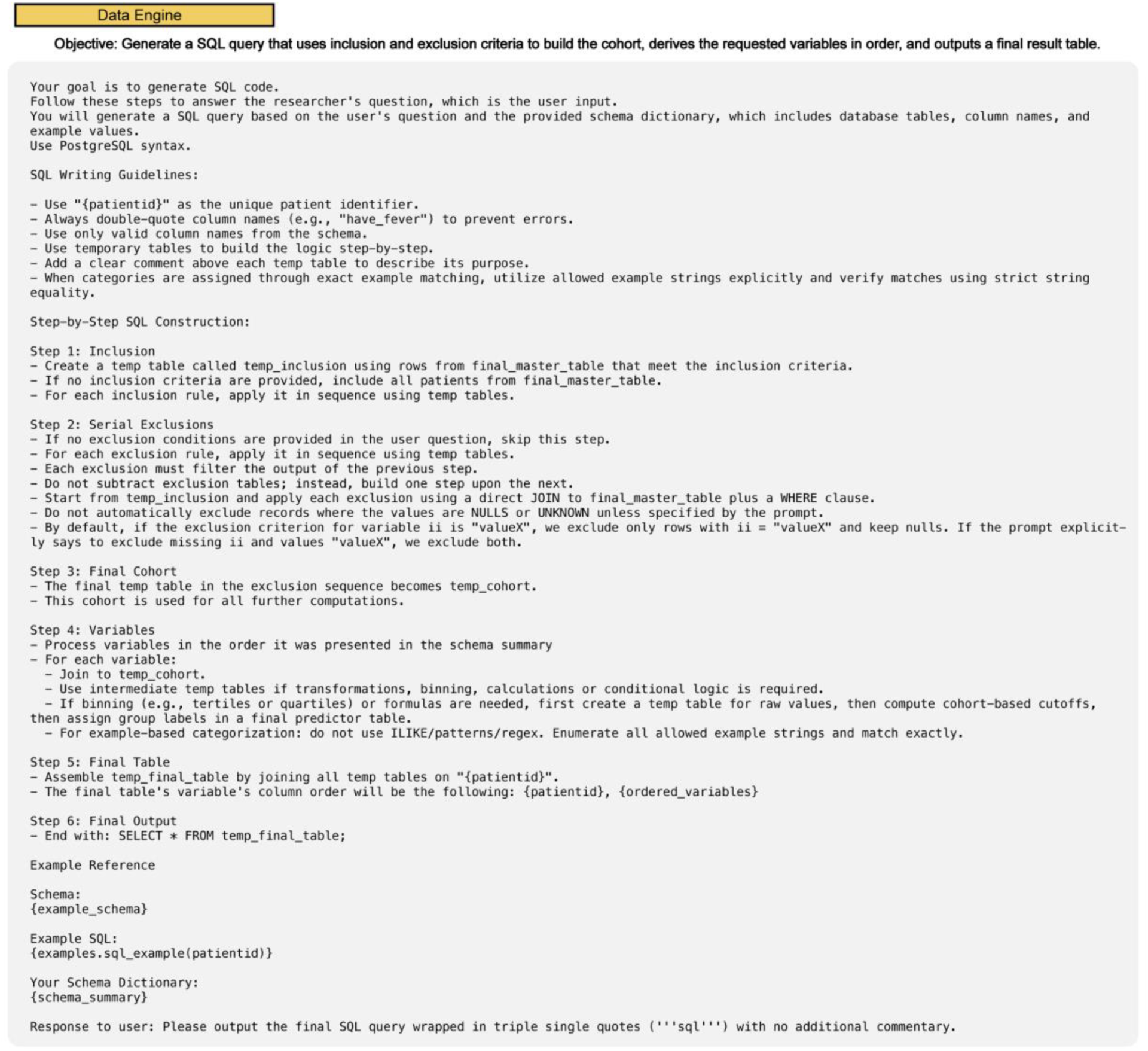
Prompt used in the Data Engine module. The LLM follows a sequence of steps to generate and format SQL queries to ensure consistency and facilitate review. A one shot example is included. The same prompt was applied across all experiments in this study.

**Supplementary Table 1.** Examples of text inputs to the LATCH framework.

| Dataset | Analysis | LATCH Input Query Example |
| --- | --- | --- |
| NHANES | Prevalence | We will get a weighted prevalence of people with diabetes. We will use NHANES data from 1999-2008. Exclude people whose age at screening is less than 20, missing any of the following visual acuity values: presenting right visual acuity, presenting left visual acuity, post-refraction right visual acuity, post-refraction left visual acuity. We will also exclude people who are missing both of the following diabetes related information: glycohemoglobin (HbA1c) (%) and self-report of a doctor's diabetes diagnosis The main predictor will be a three-level categorical variable, 'vision group', defined using the better vision eye among right and left eye: "Group 1" No VI (Reference Group): Presenting visual acuity is 20/40 or better. "Group 2" Correctable VI (CVI): Presenting acuity is worse than 20/40, but post-refraction acuity is 20/40 or better. "Group 3" Nonrefractive VI (NVI): Presenting acuity is worse than 20/40, and post refraction acuity is still worse than 20/40. Outcome is having diabetes defined as meeting one of the criteria, having glycohemoglobin (HbA1c) (%) level of 6.5 or higher, who said yes to self-report of a doctor's diabetes diagnosis, categorized as binary 1 (having diabetes) and otherwise 0. |
| AIREADI | Logistic Regression | We plan to conduct a logistic regression analysis to examine the relationship between serum albumin (g/dL) and diabetic retinopathy using NHANES data from the years 2011 to 2020. The study will include all individuals whose age in years at screening was 30 years or older who have complete data on both serum albumin (g/dL) and diabetes affected eyes/had retinopathy. Participants will be excluded if their age in years at screening is less than 30 years old, or age when first told the patient had diabetes was before age 30, or had uncertain diabetes status. Uncertainty is based on their answer to the question "Has a doctor ever told you that you have diabetes?" and if they answer as "borderline" and either have a missing glycohemoglobin (HbA1c) value or an glycohemoglobin (HbA1c) below 6.5%. We will also exclude anyone with missing data on serum albumin or diabetes affected eyes/had retinopathy. The main predictor in the analysis will be serum albumin (g/dL), categorized into quartiles (%) Q1: <25, Q2: ≥25 and <50, Q3: ≥50 and <75, Q4: ≥75. The outcome variable will be the presence or absence of diabetic retinopathy, based on responses to the question "diabetes affected eyes had retinopathy?", coded as 1 for "Yes" and 0 for "No". We will adjust the analysis for potential confounding factors including gender, age in years at screening, and race/Hispanic origin, which will be categorized as White, Black, Mexican American, or Other. |
| AIREADI | Linear Regression | We will be performing a linear regression using AIREADI data from 2023-2025. We will exclude participants with missing study group, missing values in the following variables: mesopic log contrast sensitivity od, mesopic log contrast sensitivity os, glaucoma diagnosis, cataract diagnosis, diabetic retinopathy diagnosis, AMD diagnosis, retinal vascular occlusion diagnosis. The main predictor will be a study group and covariates are age, diagnosed with glaucoma, diagnosed with cataract, diagnosed with diabetic retinopathy, diagnosed with AMD, retinal vascular occlusion stroke in the eye. The outcome will be mesopic contrast sensitivity defined as higher one among mesopic od log contrast sensitivity or mesopic os log contrast sensitivity. |
| NHANES | Cox Regression | This study aims to investigate the relationship between serum uric acid levels (mg/dl) and all-cause mortality. We will be using NHANES data between 1999 and 2018, the analysis will employ a weighted Cox proportional hazards regression model and exam weights. The study cohort will be composed of participants age in years at screening 20 years and older who meet the criteria for diabetes when an individual is considered to have diabetes if they reported doctor told them they have diabetes, had a glycohemoglobin A1c (HbA1c) level ≥ 6.5%, or a fasting plasma glucose level (mg/dl) is 126 mg/dL or greater. Individuals will be excluded from the analysis if their pregnancy exam was positive. Participants with missing laboratory values for serum uric acid (mg/dl), serum creatinine (mg/dl), or glycohemoglobin (HbA1c) (%), will be excluded. Anyone who reported ever being told they had cancer or a malignancy will be excluded, as well as those missing mortality data or months followup examination, will be excluded. The primary predictor for this analysis is the serum uric acid level (mg/dl), which will be categorized into five groups based on its distribution in the study population, using quintiles as cut-offs QN1: <20%, QN2: ≥20 and <40%, QN3: ≥40 and <60%, QN4: ≥60 and <80%, QN5: ≥80%, The main outcome is all-cause mortality, categorized as 1 or 0, with months followup examination serving as the time-to-event variable. The analysis will control factors, including age in years at screening, gender, and race/Hispanic origin, which is categorized as Non-Hispanic White, Non-Hispanic Black, Mexican American or Hispanic, and Other. |

**Supplementary Table 2.**
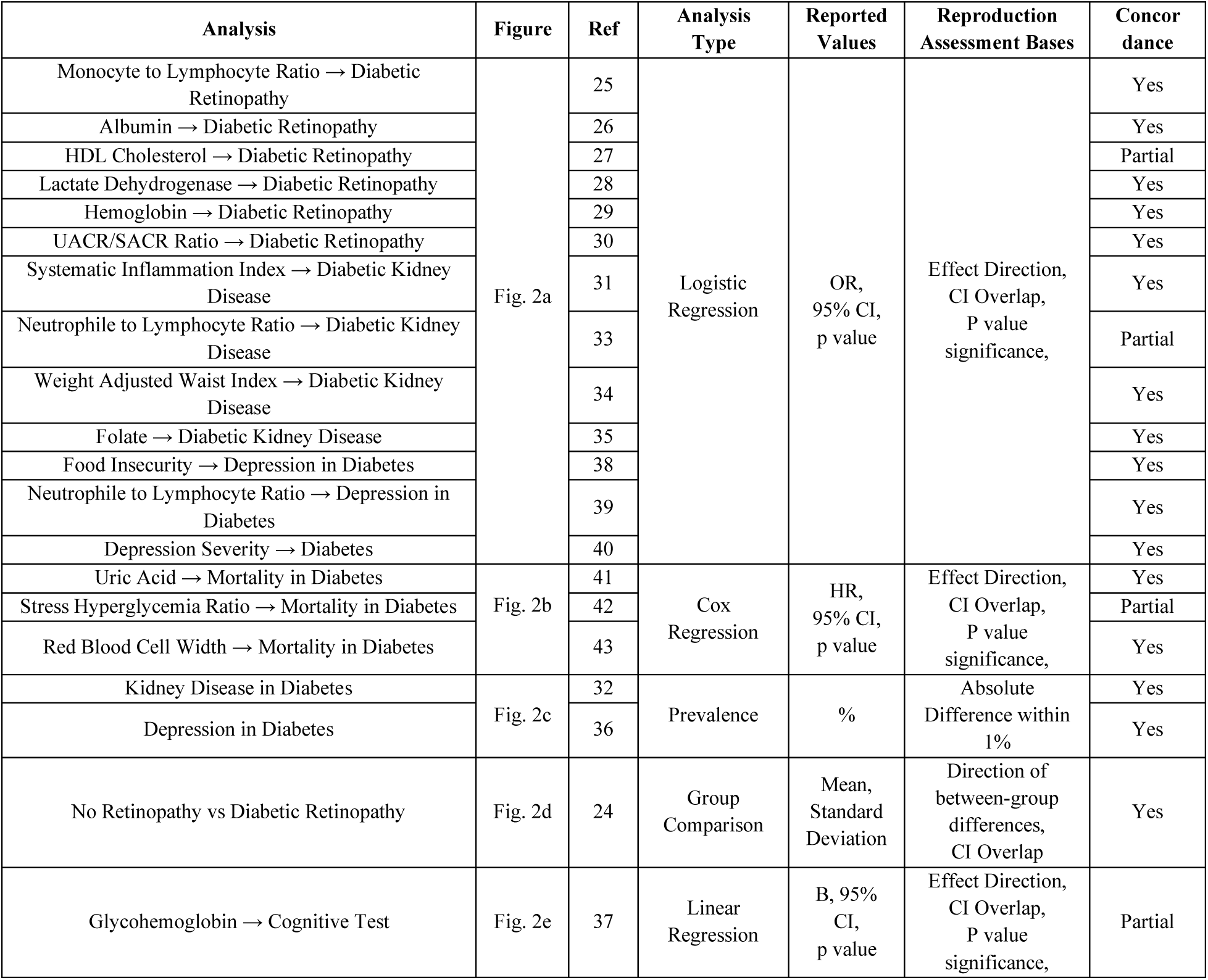
Summary of the 20 studies included in the LATCH reproduction analysis. Analysis refers to the analysis name reported in Fig. 2. The Ref column indicates the corresponding publication number listed in the references. The Figure column corresponds to the subfigures in Fig. 2 representing each reproduced analysis. Reported Values are the results originally published in each study, whereas Replication Assessment Bases are the metrics used to evaluate the extent of reproduction. Concordance indicates the overall reproduction outcome: Yes if all reproduction assessment bases were fulfilled; Partial if one or more criteria showed discrepancies; and No if none of the criteria were met.

**Supplementary Table 3.** Examples of messages generated by safeguards implemented in the Planner, Variable Mapper, and Data Engine modules.

| LATCH Module | Safeguard Activation Condition | Safeguard Message Example |
| --- | --- | --- |
| Planner | User input requires clarification due to missing, contradictory or ambiguous information | Review Required. This analysis plan is mostly well-defined, but there are a few points that need clarification to ensure the analysis can be conducted accurately: Exclusion Criteria Ambiguity: The exclusion criterion "Participants will be excluded if they are really sick and missing a lot of information" is too vague. Please specify how "really sick" will be defined (e.g., based on specific medical conditions, number of missing variables, or a particular health status indicator) and what constitutes "a lot of information" (e.g., a threshold for missing data points). |
|  |  | Review Required. The requested "machine learning analysis" is not one of the supported analysis types (logistic regression, linear regression, cox regression, group comparison, prevalence, or mediation analysis). However, based on your description of a binary outcome (presence or absence of diabetic retinopathy), a predictor (serum albumin), and several covariates, a **logistic regression** would be the most appropriate statistical analysis to examine the relationship you've described. Please confirm if a **logistic regression** would be an acceptable analysis type for your study. |
| Variable Mapper | Inconsistencies are detected in the variables matched to user-input keywords across cycles | Multiple variables candidates detected for 'A1C ≥ 6.5%' across cycles: ['glycohemoglobin', 'what_was_your_last_a1c_level'] Searching for a candidate available in all cycles...Found a candidate that is available from all cycles: 'glycohemoglobin'(From: Diabetes - diq_g) If you prefer to use other candidates, consider adjusting your time period and replace A1C ≥ 6.5% with other candidate. Rebuilding results for 'A1C ≥ 6.5%' using 'glycohemoglobin'. |
|  |  | Multiple variables candidates detected for 'renal albumin loss' across cycles: ['albumin_urine_ug_ml', 'albumin_creatinine_ratio_mg_g'] Searching for a candidate available in all cycles...Skipping candidate 'albumin_creatinine_ratio_mg_g' (From: Albumin & Creatinine - Urine - alb_cr_f). Missing from cycles: [(2007, 2008), Found a candidate that is available from all cycles: 'albumin_urine_ug_ml' (From: Albumin & Creatinine - Urine - alb_cr_f) If you prefer to use other candidates, consider adjusting your time period and replace renal albumin loss with other candidate. Rebuilding results for 'renal albumin loss' using 'albumin_urine_ug_ml'. |
| Data Engine | SQL did not run due to execution error (iterative correction up to three times) | The error "syntax error at or near "do"" occurs because `do` is a reserved keyword in PostgreSQL.In the `CREATE TEMP TABLE temp_final_table` statement, the alias `do` is used for `temp_depression_outcome`. To fix this, either quote the alias as "`do`" or change the alias to a non-reserved word (e.g., `tdo`). Replace `do.depression_outcome` with "`do`.depression_outcome"<br>... {Fixed SQL Query} |
|  |  | The error `function percentile_cont(numeric, text) does not exist` indicates that the `PERCENTILE_CONT` function is being called with a `text` type argument in its `ORDER BY` clause, but it expects a `numeric` type.<br>... {Fixed SQL Query} |

